# Design and implementation of multiplexed amplicon sequencing panels to serve genomic epidemiology of infectious disease: a malaria case study

**DOI:** 10.1101/2021.09.15.21263521

**Authors:** Emily LaVerriere, Philipp Schwabl, Manuela Carrasquilla, Aimee R. Taylor, Zachary M. Johnson, Meg Shieh, Ruchit Panchal, Timothy J. Straub, Rebecca Kuzma, Sean Watson, Caroline O. Buckee, Carolina M. Andrade, Silvia Portugal, Peter D. Crompton, Boubacar Traore, Julian C. Rayner, Vladimir Corredor, Kashana James, Horace Cox, Angela M. Early, Bronwyn L. MacInnis, Daniel E. Neafsey

## Abstract

Multiplexed PCR amplicon sequencing (AmpSeq) is an increasingly popular application for cost-effective monitoring of threatened species and managed wildlife populations, and shows strong potential for genomic epidemiology of infectious disease. AmpSeq data for infectious microbes can inform disease control in multiple ways, including measuring drug resistance marker prevalence, distinguishing imported from local cases, and determining the effectiveness of therapeutics. We describe the design and comparative evaluation of two new AmpSeq assays for *Plasmodium falciparum* malaria parasites: a four-locus panel (‘4CAST’) composed of highly diverse antigens, and a 129-locus panel (‘AMPLseq’) composed of drug resistance markers, highly diverse loci for measuring relatedness, and a locus to detect *Plasmodium vivax* co-infections. We explore the performance of each panel in various public health use cases with *in silico* simulations as well as empirical experiments. We find that the smaller 4CAST panel performs reliably across a wide range of parasitemia levels without DNA pre-amplification, and could be highly informative for evaluating the number of distinct parasite strains within samples (complexity of infection), and distinguishing recrudescent infections from new infections in therapeutic efficacy studies. The AMPLseq panel performs similarly to two existing panels of comparable size for relatedness measurement, despite differences in the data and approach used for designing each panel. Finally, we describe an R package (paneljudge) that facilitates design and comparative evaluation of AmpSeq panels for relatedness estimation, and we provide general guidance on the design and implementation of AmpSeq panels for genomic epidemiology of infectious disease.

## Introduction

Genetic data are a valuable resource for understanding the epidemiology of infectious disease. The value of this data type has been highlighted by the COVID-19 pandemic, for which viral sequence analysis has greatly informed patterns of disease spread and evolution, influencing public health policy decisions around the world (Oude Munnink et al., 2021). Applications of genetic data in epidemiology extend from viral and bacterial outbreak management (Baker, Thomson, Weill, & Holt, 2018; Coll et al., 2017; Quick et al., 2016) to the study of eukaryotic parasites underlying important diseases such as malaria, toxoplasmosis, helminthiasis, leishmaniasis and Chagas disease.

Many use cases (applications) of genetic data have been identified for malaria (Dalmat, Naughton, Kwan-Gett, Slyker, & Stuckey, 2019), the leading parasitic killer worldwide (WHO, 2019), include tracking the spread of drug/insecticide resistance genetic markers and diagnostic resistance mutations (Chenet et al., 2016; Jacob et al., 2021; Kayiba et al., 2021; Lautu-Gumal et al., 2021; Miotto et al., 2020), assessing disease transmission levels (Daniels et al., 2015; Galinsky et al., 2015), identifying sources of infections and imported cases (Liu et al., 2020; Tessema et al., 2019), and estimating genetic connectivity among different populations (Taylor et al., 2017). Malaria parasite genetic data also have demonstrated utility in therapeutic efficacy studies, for distinguishing recrudescent infections potentially indicative of low drug efficacy from reinfections or relapses from dormant liver stages (Gruenberg, Lerch, Beck, & Felger, 2019; Jones et al., 2021). In the malaria field, these applications are served by different types of genetic data produced at varying resolution, technical complexity, and cost, ranging from genetic panels that may comprise as few as 8 – 12 polymorphic microsatellites (MS) or 24 single nucleotide polymorphisms (SNPs) (Baniecki et al., 2015; Daniels et al., 2008), to whole genome sequencing (WGS) data (Miotto et al., 2015; Takala-Harrison et al., 2015).

To be scalable and sustainable, genetic data should be produced at the minimum resolution that provides robust support for the intended analysis application. Whole genome sequencing (WGS) data provide the most complete population genomic perspective on an organism of interest. However, the cost and technical challenges of generating, storing, and interpreting WGS data are impediments to scalability and widespread implementation for organisms with large genomes, or microbes with small genomes in samples dominated by host DNA. Targeted sequencing approaches that focus deep coverage on select genomic regions of interest using multiplexed PCR amplification (AmpSeq) are finding increased application in conservation genomics and fisheries biology (Baetscher, Clemento, Ng, Anderson, & Garza, 2018; Hargrove, McCane, Roth, High, & Campbell, 2021; Natesh et al., 2019; Schmidt, Campbell, Govindarajulu, Larsen, & Russello, 2020), and can serve genomic epidemiology of infectious diseases by focusing sequencing coverage on the most informative regions of pathogen or parasite genomes, instead of typically dominant host genomes (Jacob et al., 2021; Tessema et al., 2020).

Recent work on AmpSeq protocols for genotyping malaria and trypanosomatid parasites has confirmed the viability of this approach with low-parasitemia host and vector samples, where parasite DNA comprises a very small fraction of the total sample (Jacob et al., 2021; Schwabl et al., 2020; Tessema et al., 2020). Furthermore, one recent study has confirmed the value of designing amplicons to capture multi-SNP ‘microhaplotypes’, which exhibit polyallelic rather than biallelic diversity to facilitate relatedness inference (Tessema et al., 2020). New relatedness-based analytical approaches for genomic epidemiology are currently developing for malaria parasites and other sexually recombining pathogens (Henden, Lee, Mueller, Barry, & Bahlo, 2018; Schaffner, Taylor, Wong, Wirth, & Neafsey, 2018). The use of genomic data for estimation of recent common ancestry shared by pairs or clusters of parasites or mosquitoes has shown strong potential to provide epidemiologically useful inferences over very small geographic distances (10s-100s of kilometers) and short time scales (weeks to months) relative to traditional population genetic parameters of population diversity and divergence (Cerqueira et al., 2017; Taylor et al., 2017). While many analyses of recent common ancestry in malaria parasites to date have used WGS data, targeted genotyping of as few as 200 biallelic SNPs or 100 polyallelic loci (*e*.*g*., microsatellites or microhaplotypes) may also be used to infer recent common ancestry with necessary precision (Taylor, Jacob, Neafsey, & Buckee, 2019), making AmpSeq an excellent candidate to serve relatedness estimation.

However, there remains uncertainty in the molecular epidemiology field as to the suitability of existing panels for profiling parasite or pathogen populations in specific geographic locations that did not inform the original panel designs, and it is unclear which protocol features are most conducive to implementation in both high and low resource settings. Should each disease field adopt a common multiplexed amplicon protocol and panel, or should bespoke panels be implemented regionally to address genetically distinct parasite populations and specific use cases?

To address these questions, in this manuscript we describe the design and comparative evaluation of two new multiplexed amplicon assays for *Plasmodium falciparum* malaria parasites: a four-locus panel composed of highly diverse loci, ideal for estimating the number of genetically distinct strains within an infection (Complexity of Infection; COI) as well as distinguishing pre-existing vs. new infections in any geographic setting, and a 129-locus panel composed of drug resistance markers and many diverse loci for relatedness inference designed for application in South America (a region that did not inform previously published panel designs) as well as other geographic regions. Both assays use non-proprietary reagents (including standard PCR oligos) in order to maximize accessibility and affordability in malaria-endemic regions. The panels are supported by new open-source bioinformatic analysis pipelines to facilitate widespread use. We also show that the core sets of multiplexed PCR oligos can flexibly accommodate most new targets not included in the original design, allowing for panel customization towards detecting locally relevant resistance markers, polymorphic loci, and co-infecting parasite species. We use whole genome sequencing data to explore the degree to which our newly described and previously published genotyping panels can serve studies in diverse geographies, versus the alternative of customizing panels with targets that are locally informative but not globally useful. We suggest there is value in genotyping panels that can be flexibly adapted to incorporate informative targets from pathogen populations of interest. The analyses and resources described in this manuscript clarify the rapidly diversifying options for targeted microbial sequencing (**Fig. 1**), by providing tools and guidance for the comparative evaluation and refinement of AmpSeq panels.

**Figure 1.**
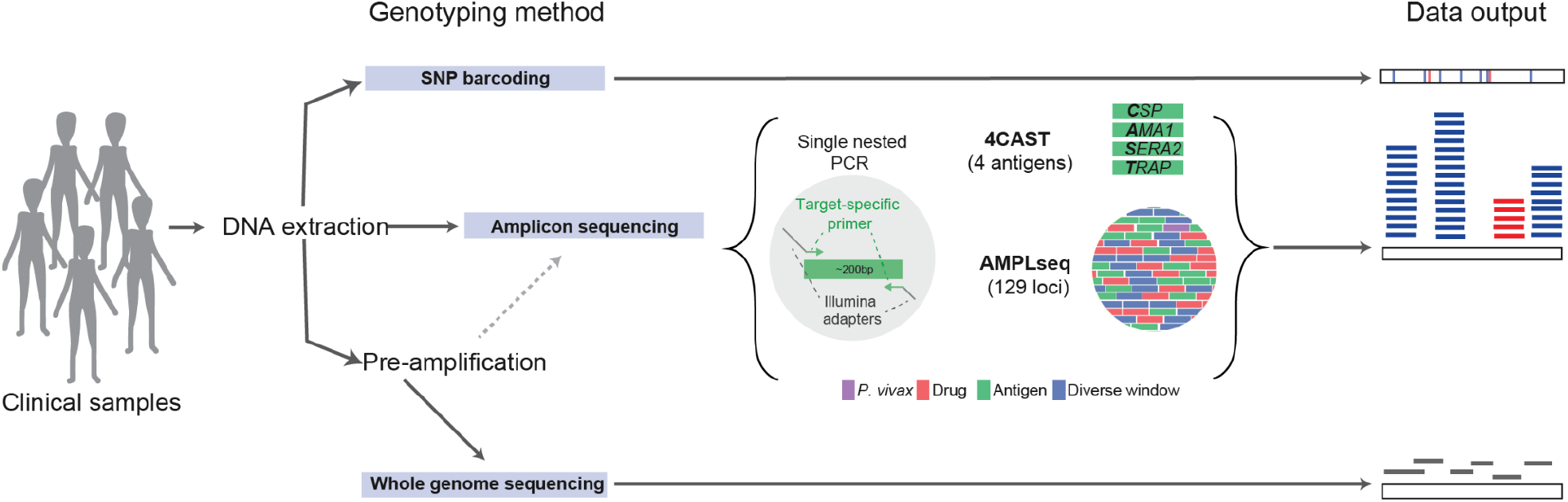
Amplicon sequencing and other genotyping approaches for genomic epidemiology of infectious diseases. Schematic of three common approaches for molecular surveillance data generation. Genomic DNA can be extracted from clinical samples and then processed using any of the three methods shown: SNP barcoding, amplicon sequencing, or whole genome sequencing (WGS). Our two amplicon panels, AMPLseq and 4CAST, are shown, with representations of their loci and amplification. Pre-amplification (selective whole genome amplification), which increases the ratio of parasite to human DNA in samples, is generally recommended for WGS and some amplicon sequencing panels (AMPLseq, but not 4CAST). SNP barcoding provides data in the form of variant calls at each SNP; amplicon sequencing provides extremely deep coverage at select, small regions of the genome; and WGS generally provides shallower coverage of the entire genome.

## Materials and Methods

### Panel designs

We developed a small multiplex of four highly polymorphic antigenic loci, dubbed ‘4CAST’: *CSP, AMA1, SERA2*, and *TRAP* (**Fig. 2**). All four amplicons use previously published primer sequences (Miller et al., 2017; Neafsey et al., 2015), as no modification was required for successful multiplexing.

**Figure 2.**
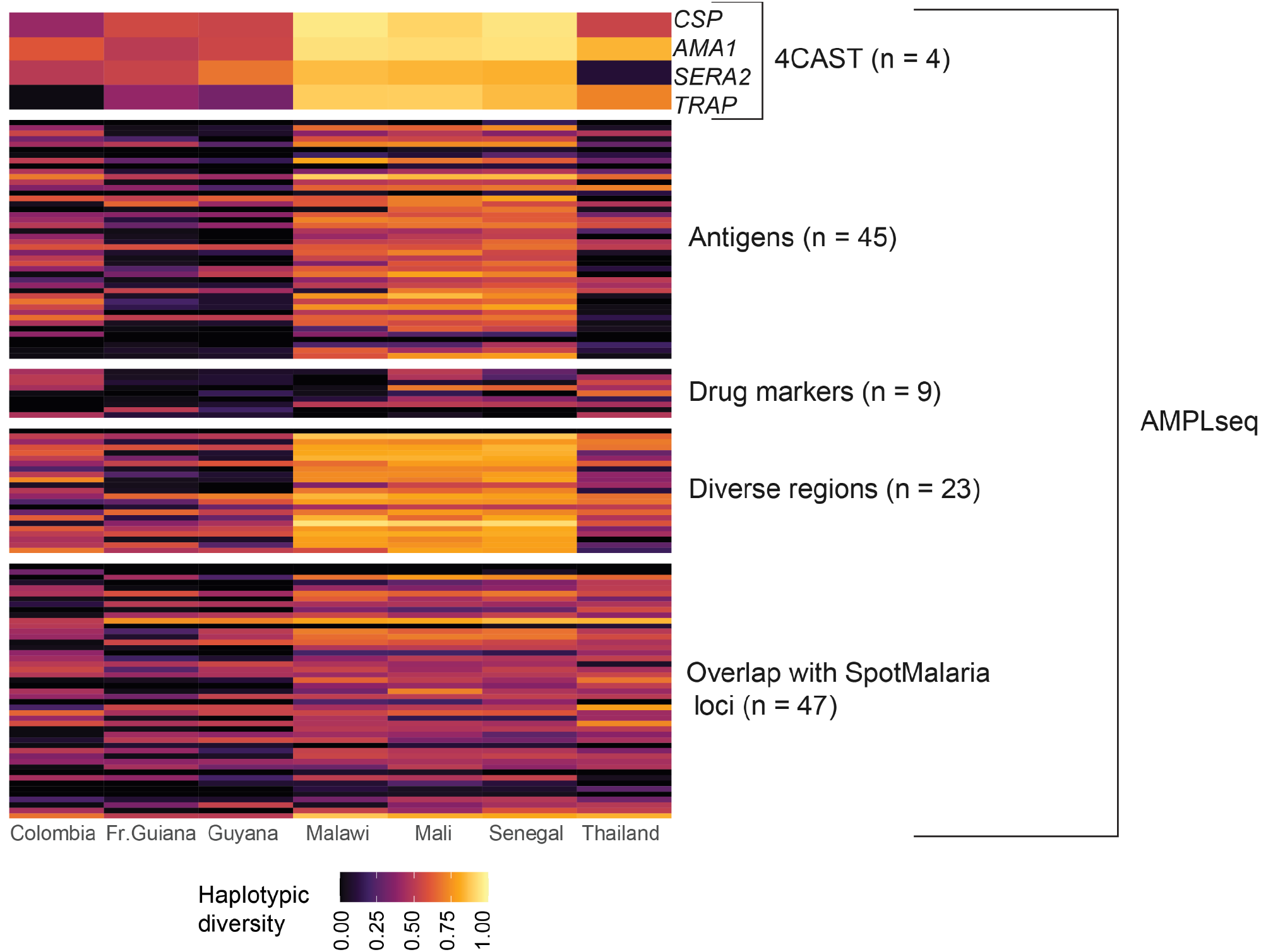
Global characterization of loci in the 4CAST and AMPLseq panels. Estimates of diversity of each locus in the 4CAST and AMPLseq panels, with one locus per row. We estimated haplotypic diversity from monoclonal *P. falciparum* WGS data from each country. The top 4 loci shown represent the 4CAST loci, which are also included in the AMPLseq panel. All 128 *P. falciparum* loci in the AMPLseq panel are shown; the single *P. vivax* locus is not shown.

In designing the larger ‘AMPLseq’ multiplexed amplicon panel, we first built a large pool of candidate loci, anticipating significant attrition of candidates due to primer incompatibility. We prioritized four classes of loci: loci within antigens of interest (Helb et al., 2015), loci with high population diversity for relatedness inference (Taylor et al., 2019), loci included in the SpotMalaria v1 panel (Chang et al., 2019; Jacob et al., 2021), and known drug resistance markers. We contracted the services of GTseek LLC (https://gtseek.com) to design multiplexed oligo panels according to the criteria previously described for the Genotyping-in-Thousands by sequencing (GT-seq) protocol (Campbell, Harmon, & Narum, 2015) (**S1 Supporting information**). We optimized the final primer set and and reaction conditions through several sequencing runs and determined that the primers for the four 4CAST loci (*CSP, AMA1, SERA2, TRAP*) could be added to the panel without impacting amplification of the other loci. We also successfully added primers amplifying known markers of antimalarial drug resistance within the genes *dhfr, dhps, mdr1*, and *kelch13* (**S2 Table**). Furthermore, we successfully added previously described primers for *PvDHFR* (Lefterova, Budvytiene, Sandlund, Färnert, & Banaei, 2015) in order to identify *P. falciparum / P. vivax* co-infections that have gone undetected in preliminary screening by microscopy or rapid diagnostic test (RDT). The final panel, dubbed ‘AMPLseq’ (short for Assorted Mix of *Plasmodium* Loci) contains this single *P. vivax* locus and 128 *P. falciparum* loci (**Fig. 2**), with a median length across all amplicons of 276 bp (**S1 Fig**.).

### Panel protocols

To create the primer pool used in 4CAST PCR1, we combined 100 µM of each 4CAST primer (**S3 Table**) and diluted the combined primer mix to 6.25 µM per primer in nuclease-free water (NF dH_2_O). Each 10.5 µl PCR1 reaction incorporated 1.5 µl combined primer mix, 5 µl KAPA HiFi HotStart ReadyMix (2x), and 4 µl sample template. PCR1 amplification consisted of an initial incubation step at 95 °C (3 min); 25 amplification cycles at 95 °C (20 s), 57 °C (15 s) and 62 °C (30 s); and a final extension step at 72 °C (1 min). Each 12.2 µl PCR2 reaction (which adds sample indices and sequencing adapters) incorporated 2.2 µl unique dual index (10 µM Illumina Nextera DNA UD Indexes), 5 µl KAPA HiFi HotStart ReadyMix (2x), 2 µl NF dH_2_O and 3 µl PCR1 product. PCR2 consisted of an initial incubation step at 95 °C (1 min); 10 amplification cycles at 95 °C (15 s), 55 °C (15 s) and 72 °C (30 s); and a final extension step at 72 °C (1 min). We combined PCR2 products in equal volumes and performed double-sided size selection using Agencourt AMPure XP beads (Beckman Coulter): we incubated 100 µl library with 55 µl beads, immobilized beads via magnet rack, and transferred the supernatant to a new tube. We incubated the transferred supernatant with 20 µl beads and washed immobilized beads twice with 80% ethanol. We eluted the library in 25 µl EB buffer (10 mM Tris-Cl, pH 8.5), subsequently adding 2.5 µl EB buffer with 1% Tween-20. We verified size selection via Agilent BioAnalyzer 2100 and sequenced the selected library at 6 pM with >10% PhiX in paired-end, 500-cycle format using MiSeq Reagent Kit v2 (**S1 Protocol**).

We followed a similar nested PCR and pooled clean-up procedure for AMPLseq library construction. Primer sequences, input volumes and concentrations are listed in **S3 Table** and PCR conditions and size selection steps are shown in **S2 Protocol**. As detailed therein, library construction for AMPLseq library construction differs in a few minor aspects. For example, primer input quantities vary slightly (800 pmol +/- 33%) to account for amplification rate differences among loci. PCR1 products are diluted 1:12 in NF dH_2_O prior to PCR2 and only single-sided (left-tailed) bead-based size selection is used to enhance yield. Sequencing also occurs via paired-end, 500-cycle MiSeq but with a higher final library loading concentration (12 pM) and a lower fraction of PhiX (8%).

### Mock samples

We generated mock samples from parasite lines 3D7 and Dd2, cultured at 3% hematocrit in commercially obtained red blood cells as previously described (Trager & Jensen, 1976). We extracted genomic DNA using the Qiagen Blood and Tissue Kit on cells previously lysed with 0.15% saponin. We generated positive control template representing DNA extractions from whole human blood infected with 10,000 monoclonal 3D7 parasites/µl by diluting genomic DNA from 3D7 to 0.25 ng/µl in 10 ng/µl human genomic DNA, and storing in 10 mM Tris-HCl (pH 8.0), 1 mM EDTA (Promega, Madison, WI). We generated further control templates representing 1000, 100, and 10 3D7 parasites/µl by serial 1:10 dilution of the 10,000 3D7 parasites/µl control, likewise using 10 ng/µl human genomic DNA as diluent. We also generated a 10,000 parasites/µl positive control as described above but using Dd2 instead of 3D7 strain genomic DNA. We generated mixed-strain control templates by combining the 10,000 3D7 parasites/µl control with this 10,000 Dd2 parasites/µl control at 1:1, 3:1, and 10:1 ratios (respectively). We serially diluted the 1:1 ratio to 1000, 100, and 10 parasites/µl concentrations and diluted the 3:1 and 10:1 ratios to 1000 and 100 parasites/µl concentrations using 10 ng/µl human genomic DNA diluent as before. We also applied selective whole genome amplification (sWGA) to all above control templates representing ≤ 1000 parasites/µl. The 50 µl sWGA reaction followed Oyola *et al*. 2016 (Oyola et al., 2016) with the exception of fixing template input volume to 10 µL. We purified sWGA products with Agencourt AMPure XP beads (Beckman Coulter) on the KingFisher Flex (**S3 Protocol**) and verified amplification success via NanoDrop (ThermoFisher Scientific).

### Clinical samples

We tested the panels on clinical dried blood spot (DBS) samples from Mali and Guyana. Tran *et al*. collected samples in Kalifabougou, Mali in 2011 – 2013 as previously described (Tran et al., 2013). The Kalifabougou cohort study was approved by the Ethics Committee of the Faculty of Medicine, Pharmacy and Dentistry at the University of Sciences, Technique and Technology of Bamako, and the Institutional Review Board of the National Institute of Allergy and Infectious Diseases, National Institutes of Health (NIH IRB protocol number: 11IN126; https://clinicaltrials.gov/; trial number NCT01322581). Written informed consent was obtained from participants or parents or guardians of participating children before inclusion in the study. The Guyana Ministry of Health collected samples from Port Kaituma and Georgetown, Guyana between May – August 2020, by spotting participants’ whole blood onto Whatman FTA cards and storing the samples with individual desiccant packets at room temperature. Informed consent (or parental assent for minors) was obtained for all subjects according to protocols approved by ethical committees.

We punched DBS samples 3 – 5 times into a 96-well deep well plate using the DBS pneumatic card puncher (Analytical Sales and Services, Inc.) equipped with a 3 mm cutter. We then extracted gDNA following the DNA purification from buccal swab section of the KingFisher Ready DNA Ultra 2.0 Prefilled Plates for KingFisher Flex instruments protocol (ThermoFisher Scientific) with minor modifications (**S4 Protocol**). We used the same sWGA procedure as above on the extracted gDNA.

### Whole genome sequencing and variant calling

We performed whole genome sequencing on clinical samples collected in Guyana to validate 4CAST and AMPLseq outcomes. We performed sWGA on DNA samples as described above to enrich parasite DNA. We used the enriched DNA to construct Illumina sequencing libraries from the amplified material using the NEBNext Ultra II FS DNA prep kit (NEB #E6177) prior to sequencing on an Illumina HiSeqX instrument at the Broad Institute, using 150 bp paired-end reads, targeting a sequencing depth of at least 50X coverage. We aligned reads to the *P. falciparum* v3 reference genome assembly using BWA-MEM (Li, 2013) and called SNPs and INDELs using the GATK HaplotypeCaller (DePristo et al., 2011; McKenna et al., 2010; Van der Auwera et al., 2013) according to the best practices for *P. falciparum* as determined by the Pf3k consortium (https://www.malariagen.net/resource/34). Analyses were limited to the callable segments of the genome (Miles et al., 2016) and excluded sites where over 20% of samples were multiallelic. Data from these samples were submitted to the NCBI Sequence Read Archive (http://www.ncbi.nlm.nih.gov/sra) under accession PRJNA758191.

### Amplicon data analysis

We developed an application named AmpSeQC (**S2 Supporting information**) to assess sequence quality and amplicon/sequence run success (**S2 Fig**.). We also used AmpSeQC for *P. vivax* detection by applying a concatenation of the *P. falciparum* 3D7 and *P. vivax* PvP01 reference genomes during the BWA-MEM alignment step. For in-depth assessment of *P. falciparum* sequence variation, we processed paired-end Illumina sequencing data in the form of FASTQ files using a custom analysis pipeline (**S2 Supporting information**) that leverages the Divisive Amplicon Denoising Algorithm (DADA2) tool designed by Callahan *et al*. 2016 (Callahan et al., 2016) to obtain microhaplotypes (**S2 Fig**.). We mapped microhaplotypes obtained from DADA2 against a custom-built database of 3D7 and Dd2 reference sequences for each amplicon locus and filtered microhaplotypes based on edit distance, length, and chimeric identification, using a custom R script. (**S2 Supporting information**). We summarized observed sequence polymorphism into a concise format by converting individual microhaplotypes into pseudo-CIGAR strings using a custom python script. Microhaplotypes were discarded if supported by fewer than 10 read-pairs or by less than 1% total read-pairs within the locus, or if they exhibited other error features (**S3 Supporting information**).

We analyzed native and pre-amplified mock samples to determine precision and sensitivity of the DADA2 pipeline and filters. We defined a true positive (TP) as a microhaplotype with a pseudo-CIGAR string identical to the reference strain (either 3D7 or Dd2). We defined a false positive (FP) as a microhaplotype with a pseudo-CIGAR string not matching 3D7 (in the case of samples containing only 3D7) or not matching 3D7 or Dd2 (in the case of the mixes), and we defined a false negative (FN) as a locus without any correct microhaplotype representation. We defined precision as TP/(TP+FP), and sensitivity (recall) as TP/(TP+FN). Forty-two of 128 *P. falciparum* loci in AMPLseq exhibit identical 3D7 and Dd2 reference sequences; we only included these in precision and sensitivity calculations for pure 3D7 controls (*i*.*e*., TP+FN = 128); precision and sensitivity calculations for strain mixtures considered only the 86 loci that differ between 3D7 and Dd2 reference sequences (*i*.*e*., TP+FN = 86).

All amplicon sequencing data were submitted to the NCBI Sequence Read Archive (http://www.ncbi.nlm.nih.gov/sra) under accession PRJNA758191.

### Comparator Panels

We compared AMPLseq and 4CAST to two previously published AmpSeq panels for malaria molecular surveillance, Paragon HeOME v1 (Tessema et al., 2020) and SpotMalaria v2 (Jacob et al., 2021).

Paragon HeOME v1, designed via CleanPlex algorithm (Paragon Genomics Inc, USA), contains 100 primer pairs in a single pool. These target 89 *P. falciparum* loci selected based on high diversity and differentiation (Jost D ≥ 0.21) among clinical isolates from Africa as well as eleven drug resistance-associated loci. A distinctive feature of HeOME library construction involves its requirement for bead-based clean-up and CleanPlex digestion of each sample between PCR1 and PCR2. The protocol therefore does not require sWGA prior to PCR1.

SpotMalaria v2, designed via Agena BioScience and MPrimer design software, contains 136 primer pairs divided into three different pools. These target loci were considered best able to recapitulate pairwise genetic distance, population differentiation, and sample heterozygosity inferences from global WGS data (MalariaGEN *Plasmodium falciparum* Community Project, 2016). Primers also target various drug resistance-associated loci (some known to exhibit copy number variation) and mitochondrial loci with conserved primer binding sites among *Plasmodium* spp. Library construction requires sWGA prior to PCR1 but no special processing between PCR1 and PCR2.

We also compared our amplicon panels to a 24-SNP molecular barcode assay (Daniels et al., 2008). The SNPs targeted by this Taqman qPCR-based assay were chosen principally for their high minor allele frequency (average MAF > 0.35) in parasite sample collections from Thailand and Senegal (Daniels et al., 2008).

### paneljudge and *in silico* data simulations

We used WGS data to simulate genotypic panel data for simulations. This publication uses data from the MalariaGEN *Plasmodium falciparum* Community Project as described online pending publication and public release of dataset Pf7 (https://www.malariagen.net/resource/34). Specifically, we used genomic data from monoclonal samples collected in Mali, Malawi, Senegal, and Thailand (Zhu et al., 2019), and from Colombia and Venezuela (ENA accession numbers in **S4 Table**). We also used previously published monoclonal genomic data from Guyana (SRA BioProject PRJNA543530) (Mathieu et al., 2020) and French Guiana (SRA BioProject PRJNA242182) (Pelleau et al., 2015). We used the *scikit-allel* library (Miles et al., 2020) to process the data and then estimate microhaplotype frequency and diversity. Specifically, we used the read_vcf, is_het, and haploidify_samples functions as described (**S1 Supporting information**), and we estimated haplotype frequencies with the distinct_frequencies function.

We assessed the performance of different panels for relatedness inference using simulated data. We generated data on pairs of haploid genotypes (equivalent to pairs of monoclonal malaria samples) using paneljudge (Taylor & Jacob, 2020), an R package that we built to simulate data under a hidden Markov model (HMM) (Taylor et al., 2019) (**S2 Supporting information**). For each panel, we calculated inter-locus distances from the median nucleotide position of each locus and set distances as infinite between chromosomes. For each panel and population of interest, we calculated haplotype frequency estimates using *scikit-allel*, as described above. Given these distances and frequency estimates, we simulated data using relatedness parameter values of 0.01 (unrelated), 0.50 (siblings), and 0.99 (clonal), and switch rate parameter values of 1, 5, 10, and 50. For each combination of panel, population, relatedness parameter, and switch rate parameter, we simulated data on 100 haploid genotype pairs. For each haploid genotype pair, we then generated estimates of the relatedness parameter and the switch rate parameter using paneljudge, with 95% confidence intervals (CIs), under the same model used to simulate the data. We next performed relationship classification from these estimates and CIs. For estimates of unrelated pairs (relatedness parameter of 0.01), we generously classified estimates as correct if the lower limit of the 95% confidence interval (LCI) was below or equal to 0.01 and the upper limit of the 95% confidence interval (UCI) was below 0.99. We classified estimates of sibling-level relatedness (0.50) as correct if the LCI was above 0.01 and the UCI was below 0.99. We classified estimates of clonal pairs (0.99) as correct if the LCI was above 0.01 and the UCI was above or equal to 0.99. In all relatedness levels, if the 95% confidence interval spanned both 0.01 and 0.99 (*i*.*e*., LCI < 0.01 and UCI > 0.99), then we denoted the estimate as unclassified.

To evaluate panel performance in COI estimation, we combined monoclonal WGS data to engineer *in silico* polyclonal samples using vcftools (Danecek et al., 2011). We then estimated microhaplotype frequencies for each locus of a given panel, using *scikit-allel* as described above, and counted the number of distinct microhaplotypes observed at each locus per sample. We estimated COI as the maximum number of distinct microhaplotypes observed at any locus within a sample.

To evaluate panel performance for geographic attribution, we identified microhaplotypes at loci as described above. We used the microhaplotype sequences themselves and visualized these data using the Rtsne package (Krijthe, 2015), with 5000 iterations, Θ of 0.0, and perplexity parameters of 10 (for 4CAST and the 24 SNP barcode) or 30 (for the remaining panels).

## Results

### 4CAST and AMPLseq validation

We validated assay precision (defined as TP/(TP+FP)), sensitivity (defined as TP/(TP+FN)), and depth of coverage using 3D7 mock clinical samples representing parasitemia levels between 10 and 10000 parasites/µl in 10 ng/µl human DNA. Both 4CAST and AMPLseq generated 3D7 microhaplotype calls with 100% precision for all parasitemia levels assessed, both with and without pre-amplification by sWGA. 4CAST achieved high sensitivity and depth without preliminary sWGA, generating a median of 43 read-pairs per locus from native templates representing 10 parasites/µl (**Fig. 3A**). Median depth increased to 443 and 1312.5 read-pairs per locus for native templates representing 100 and 1000 parasites/µl, respectively. Read-pair counts were also evenly distributed among 4CAST loci using native DNA (**Fig. 3A**).

**Figure 3.**
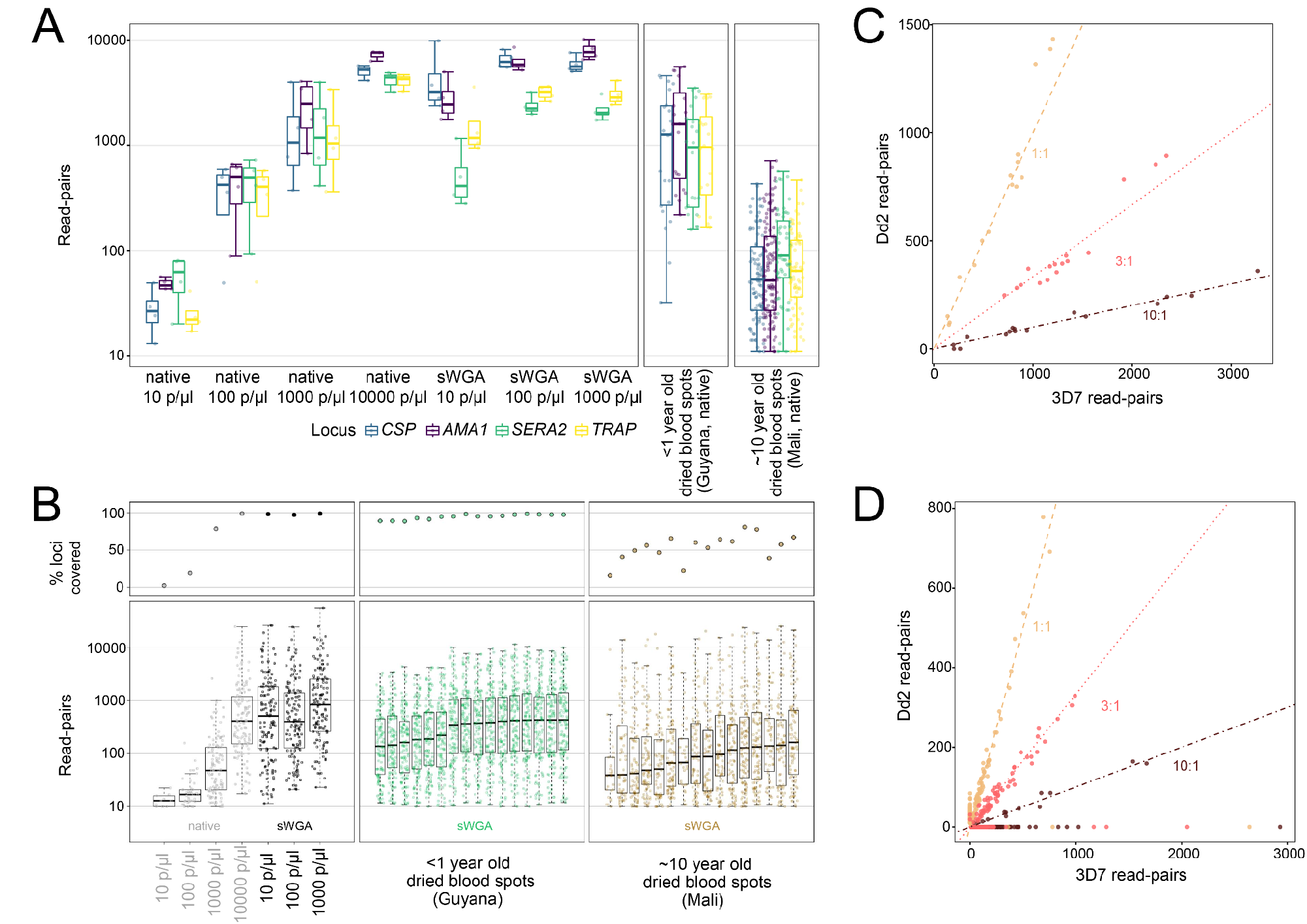
4CAST and AMPLseq panel validation with mock and clinical samples. A) Boxplots of read-pairs per locus in the 4CAST panel. The first facet shows read-depth per locus across mock samples ranging from 10 - 10000 p/µl, both native DNA and sWGA DNA (n=4 per condition). The second and third facets show read-depth per locus across two sets of clinical samples, <1 year old and ∼10 year old dried blood spots, respectively (n=32 per sample set). B) The same 3D7 mock sample sets were used to assess AMPLseq sensitivity (top left panel of B) and sequencing depth (bottom left panel of B). Each point in the bottom left panel of B represents read-pair support for one AMPLseq locus. Positions on the y-axis indicate median read-pair support across replicate samples. Low sensitivity observed using native templates (grey) representing ≤100 3D7 parasites/µl suggests that clinical samples should be pre-amplified with sWGA (results at right). C) Ratio of read-pairs from microhaplotypes assigned to 3D7 (x-axis) or Dd2 (y-axis) from mock mixtures of these DNAs in ratios of 1:1 (tan), 3:1 (pink), and 10:1 (dark red). All samples contained 1000 parasites per µl total, across both DNA sources. Dashed lines represent the expected ratio, and each point represents a 4CAST locus per sample (n=4 per condition). D) AMPLseq read-pair ratios observed in native 3D7+Dd2 mock mixtures (1000 parasites/µl) are plotted as above (C) for 4CAST.

Unlike 4CAST, AMPLseq required sWGA for 3D7 mock samples representing ≤ 100 parasites/µl (**Fig. 3B**). Following sWGA on mock samples representing 10 parasites/µl, the assay generated ≥ 10 read-pairs at a median of 126 loci, with a median of 465 read-pairs after excluding loci with fewer than 10 reads. Values were statistically similar for pre-amplified samples representing 100 parasites/µl and increased to 692 read-pairs for pre-amplified samples representing 1000 parasites/µl (**S3 Fig**.).

We also validated the sensitivity of 4CAST and AMPLseq for genotyping polyclonal infections by using mock samples containing both 3D7 and Dd2 templates (likewise in 10 ng/µl human DNA). These mixtures featured Dd2 at 50% (*i*.*e*., 1:1 3D7:Dd2 ratio), 25% (3:1), and 9% (10:1) relative abundance. Total parasitemia levels ranged between 10 and 10000 parasites/µl. Both 4CAST and AMPLseq generated microhaplotype calls with 100% precision at the 86 loci that are dimorphic between the 3D7 and Dd2 references (including all four 4CAST loci and an additional 82 loci in AMPLseq). This perfect precision was observed at all parasitemia levels in both native and pre-amplified mock mixtures of the two strains.

4CAST showed high sensitivity for Dd2 without the need for sWGA. At 1000 parasites/µl, the assay detected Dd2-specific microhaplotypes at each of its four loci in all 1:1, 3:1, and 10:1 mixture replicates (**Fig. 3C**). At 100 parasites/µl, median Dd2 sensitivity remained 100% at 1:1 and 3:1 ratios but lowered slightly to 94% at 10:1. Sensitivity for each strain decreased at 10 parasites/µl, with 1:1 ratios yielding a median of 3 target loci for 3D7 and a median of 2 targets for Dd2; median sensitivity in these samples rose to 3.5 and 3 loci (respectively) following pre-amplification with sWGA, but this led to unbalanced read-pair support between the two strains (**S4A Fig**.), possibly due to differential sWGA success on low-quality Dd2 vs. high-quality 3D7 templates. 4CAST read-pair ratios generated from native templates, by contrast, showed a very high correlation with input ratios at 100 p/µl (**S4A Fig**.) and 1000 p/µl (**Fig. 3C**). Ratios became less informative at 10 parasites/µl (**S4A Fig**.).

AMPLseq (with sWGA) was also successful in detecting Dd2-specific microhaplotypes, but only at a maximum of 77 of 86 dimorphic loci (in the 1:1 ratio at 10000 parasites/µl). Dd2-specific sequences were detected at a minimum of two dimorphic loci for all three input ratios (1:1, 3:1, 10:1) and parasitemia levels (≥ 10 p/µl) assessed. Like with 4CAST, however, the use of sWGA decorrelated read-pair ratios from input ratios (**S4B Fig**.). Dd2 sensitivity was also reduced relative to 3D7 sensitivity (median Dd2 sensitivity / 3D7 sensitivity = 58%) with sWGA. These discrepancies were not observed with native templates (**Fig. 3D**) at ≥1000 parasites/µl for which AMPLseq achieves high read-pair support without the use of sWGA.

We also tested both panels on genomic DNA extracted from dried blood spots collected by the Guyana Ministry of Health in 2020 from individuals diagnosed as *P. falciparum*-positive via a rapid diagnostic test (RDT). Ten Guyanese samples were tested with both panels, and an additional six were tested with AMPLseq. Using 4CAST, we observed coverage across all loci in all samples, with a median read-pair depth per locus of 1162 read-pairs without sWGA (**Fig. 3A**). Using AMPLseq (with sWGA), we observed a median of 122 loci with ≥10 read-pairs and a median read-pair depth of 298 read-pairs per covered locus (**Fig. 3B**).

Additionally, we tested both panels on gDNA extracted from 16 dried blood spot samples collected in Mali in 2011 (Tran et al., 2013) and subsequently stored at room temperature for ten years. Using 4CAST (without sWGA), we observed a median read-depth of 407 read-pairs per locus (**Fig. 3A**). Using AMPLseq (with sWGA), we observed a median of 75 loci with ≥10 read-pairs and a median read-depth of 112 read-pairs per covered locus (**Fig. 3B**).

### Evaluation of panel performance for relatedness

We used the R package paneljudge to assess *in silico* the impact of choosing a specific genotyping panel for relatedness inference. Considering the choice of panel, we evaluated relatedness estimation from data simulated on our 4CAST and AMPLseq panels, the SpotMalaria v2 (Jacob et al., 2021) and Paragon HeOME v1 (Tessema et al., 2020) amplicon panels, and a barcode of 24 SNPs. When data were simulated using microhaplotype frequency estimates of Senegalese parasites, we found that almost all estimates of unrelated or clonal pairs were correctly classified, regardless of the panel (**Fig. 4A)**. All three large panels also performed similarly well in accurately identifying partially-related parasite pairs, despite being the product of three distinct design processes. Neither 4CAST nor the 24 SNP barcode estimated relatedness for partially-related samples as well as the larger panels. We also evaluated panel performance in less diverse parasite populations (Colombia and Thailand), including a population not used in the panel designs (Colombia). We repeated the simulations using microhaplotype frequencies estimated with these data. Again, we found that all panels performed well for estimating relatedness of clonal pairs, and that the 24 SNP barcode and 4CAST were less likely to have correctly classified estimates of non-clonal pairs. With the data simulated using Colombian microhaplotype frequencies from the Pacific Coast region, all three large panels performed well for all three relatedness values, despite the Colombian data not having informed the design of any of the panels.

**Figure 4.**
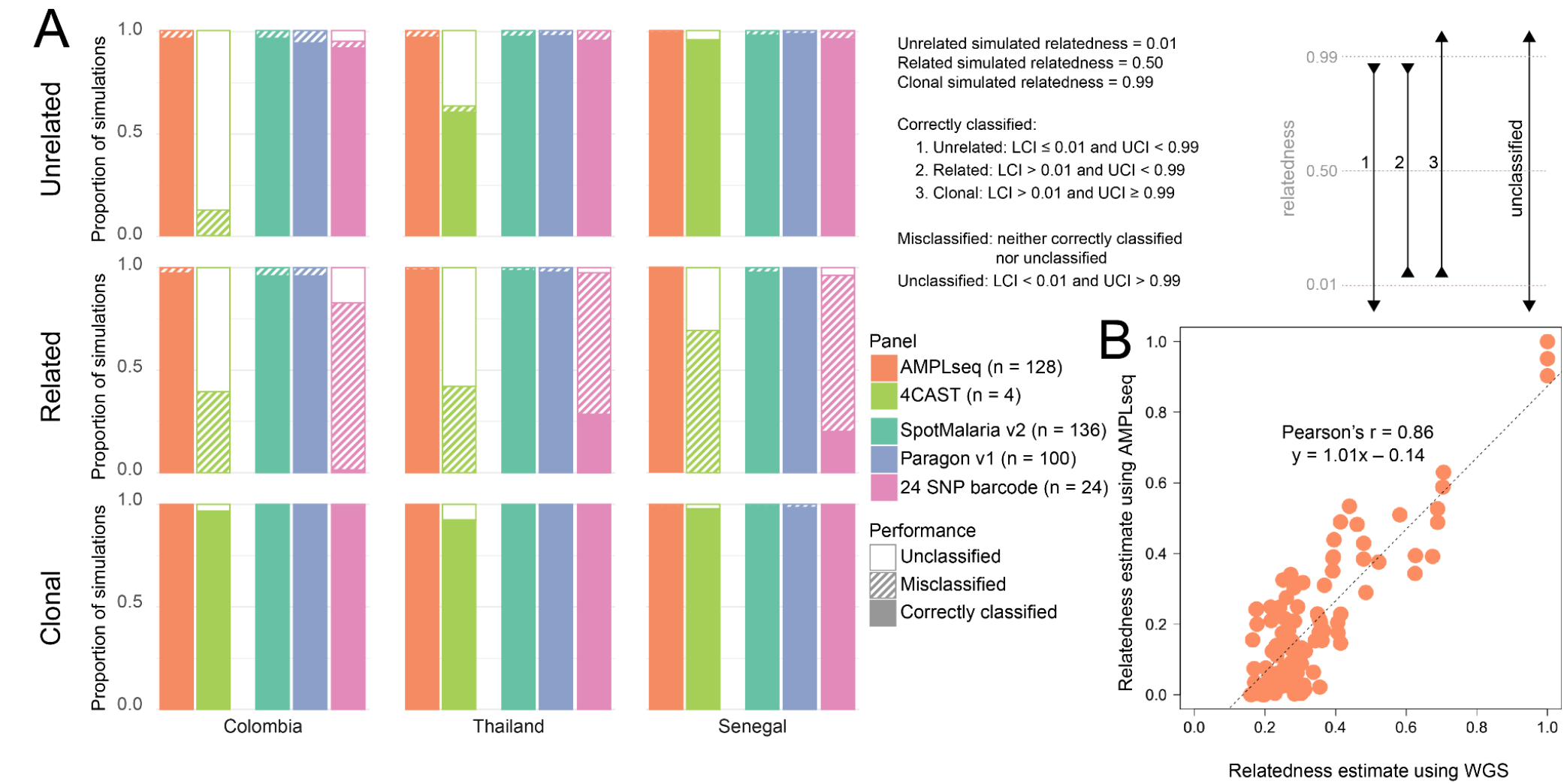
*In* silico relatedness estimation comparisons among panels and empirical AMPLseq validation against WGS. A) Evaluation of relatedness estimation from data simulated on genotyping panels using the paneljudge R package. Pairs of haploid genotypes were simulated at each locus of a panel, using microhaplotype frequencies estimated from a given parasite population (Colombia, Thailand, or Senegal, as shown in columns from left to right). Genotype pairs were simulated at three levels of relatedness: unrelated (relatedness = 0.01), related (relatedness = 0.50), and clonal (relatedness = 0.99), as shown in the rows from top to bottom. Relatedness estimates of these pairs were classified using their 95% confidence intervals (LCI = lower limit of the 95% confidence interval, UCI = upper limit of the 95% confidence interval). Estimates could be correctly classified, misclassified, or unclassified, as described in the grey box. Each bar represents the proportion of simulations per condition (n = 400) classified in each category. Bars that are filled with a color represent correctly classified simulations, bars that are hashed represent misclassified simulations, and bars that are filled with white represent simulations that were unable to be classified. The colors of the bars represent the panel used in that set of simulations. B) Empirical AMPLseq results recapitulate WGS-based relatedness inference. Points represent relatedness estimates (hmmIBD (Schaffner et al., 2018) ‘fract_sites_IBD’ computed under default settings) for pairs of Guyanese samples using WGS (n= 9408 variants) vs. AMPLseq (n = 220 variants, from within 128 AMPLseq *P. falciparum* loci).

Pairwise relatedness estimates (Schaffner et al., 2018) from AMPLseq correlated highly with those from WGS data available for the Guyanese sample set (Pearson’s r = 0.86, slope = 1.01, p < 0.001) (**Fig. 4B**). Despite patient travel history metadata suggesting infections to have occurred in various geographic regions of Guyana (**S1 Table**), AMPLseq relatedness estimates for the Guyanese sample set are significantly higher than those for the Malian sample set (Mann Whitney U, p < 0.001), consistent with anticipated lower transmission levels in Guyana. Nevertheless, the wide range of relatedness estimates (0.007 – 1) observed among Guyanese sample comparisons suggests AMPLseq capacity to indicate epidemiologically relevant microstructure even in relatively unstructured parasite populations. For example, the first (lowest) quartile of pairwise relatedness estimates from Guyana was enriched for comparisons involving A2-GUY and C5-GUY, two highly related samples that in whole genome analysis show 50% relatedness with a sample from Venezuela (SPT26229, see **S4 Table**).

### Geographic attribution

We again engineered amplicon data *in silico* to evaluate the relative signal in genotyping panels for geographic attribution of samples (**Fig. 5**). By sub-sampling WGS variant calls, calling microhaplotypes, and visualizing these data using t-SNE plots, we found that results from both the 24 SNP barcode (Daniels et al., 2008) and 4CAST distinguished samples by continent of origin, though not by country. Results from all three larger panels additionally distinguished non-African samples by country, and these panels separated East African (Malawi) from West African samples (Mali/Senegal) to varying degrees; no panel was able to distinguish between Malian and Senegalese samples in this visualization. We also added empirical AMPLseq data from 5 Guyanese samples (C3-GUY, C4-GUY, C5-GUY, C7-GUY, and C8-GUY) and WGS data sub-sampled to AMPLseq coordinates for Venezuelan sample SPT26229 (**S5 Fig**.). The AMPLseq samples formed a small cluster beside the WGS-based Guyanese and French Guianese samples. The Venezuelan sample SPT26229 also placed on the perimeter of the Guyana/French Guiana sample cluster, sharing the same axis-2 position as the empirical AMPLseq points. Results show that empirical AMPLseq data can distinguish autochthonous samples from the Guiana shield, and we expect geographic attribution in the region to improve as more data are collected from infections originating in Venezuela and other undersampled localities.

**Figure 5.**
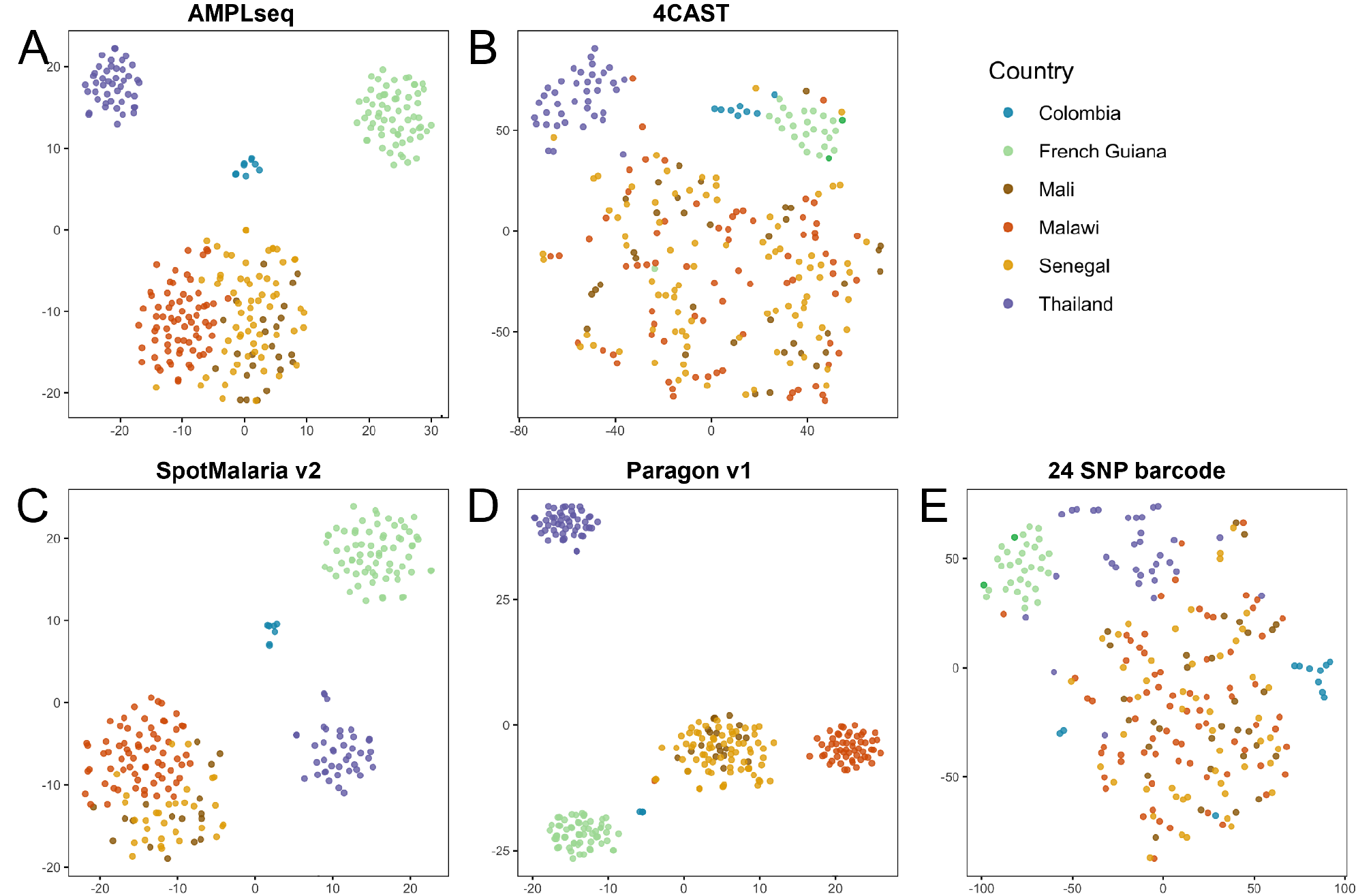
*In silico* geographic attribution comparison among panels. Visualization of WGS data subset to coordinates of genotyping panels. Microhaplotypes called at each locus were visualized using tSNE through the Rtsne package, with parameter Θ of 0.0, 5000 iterations, and a perplexity parameter of 30 (A, C, D) or 10 (B, E). Each dot represents a single sample, colored by its country of origin (which was not included in the tSNE algorithm). One genotyping panel is visualized in each plot: (A) AMPLseq, (B) 4CAST, (C) SpotMalaria v2, (D) Paragon v1, (E) 24 SNP barcode.

### COI estimation

We evaluated COI estimation based on 4CAST as opposed to the single locus *AMA1*, which is commonly used for this purpose, alone or with another locus (Lerch et al., 2017; Miller et al., 2017; Nelson et al., 2019). We engineered *in silico* samples with COI ranging from 2 – 10 (100 simulations per COI level) and used the maximum number of unique microhaplotypes present at any locus as a simple objective method to estimate COI. 4CAST provided more accurate estimates of COI than *AMA1* alone in these simulated data, especially at simulated COI levels between 5 and 7. (Fig. 6A). S6 Fig. indicates that estimation improves at simulated COI=8 using AMPLseq, but to reap the full benefit of the larger panel in practice will require a more elaborate probabilistic algorithm that accounts for variable coverage/sensitivity among loci and incorporates a multinomial approach for polyallelic loci.

**Figure 6.**
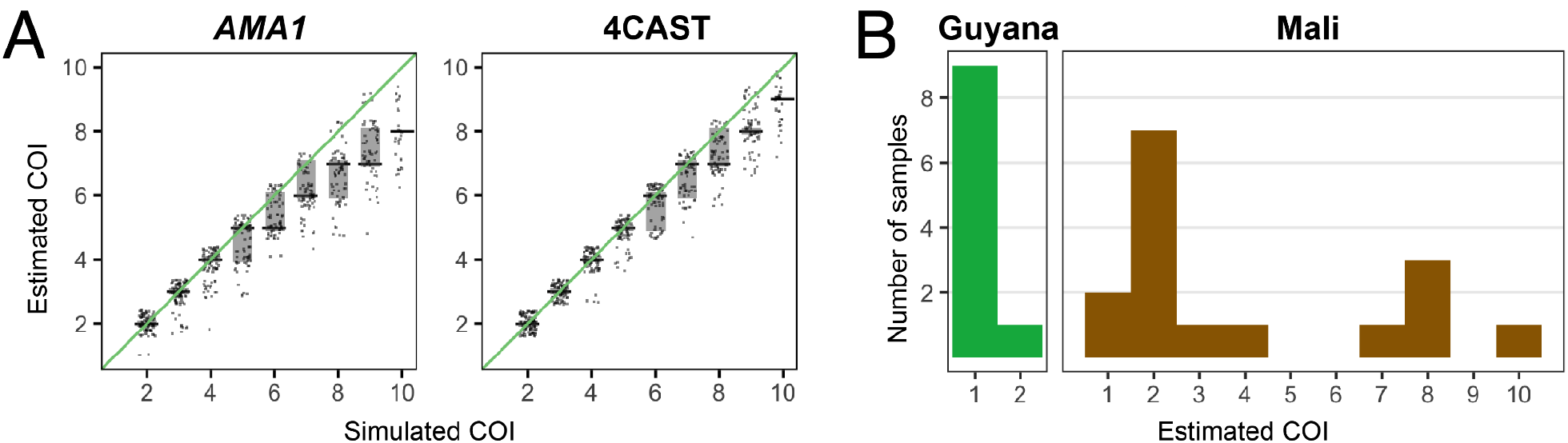
*In silico* and empirical complexity of infection (COI) inference. A) Scatter plots of estimated COI for samples simulated from combinations of monoclonal WGS data, subsetted to the loci of interest (*AMA1* locus or 4CAST loci). The x-axis represents the number of monoclonal genomes combined into each simulation, and the y-axis represents the COI estimated using the simulated data. COI was naively estimated as the maximum number of unique microhaplotypes present at any locus per sample (n=100 samples per condition). Each dot represents a sample, jittered for visibility. The black bars represent the median and light grey boxes represent the 25th – 75th quantiles. B) Estimated COI for clinical samples sequenced using 4CAST. COI was again estimated as the maximum number of unique microhaplotypes present at any locus in the sample.

In the absence of such an algorithm, we proceeded with the naive estimation method above to classify COI in the Mali and Guyana clinical samples. Only a single polyclonal infection (C6-GUY) occurred among Guyanese samples assayed by 4CAST. The repeated detection of two *CSP* and *SERA2* alternate alleles at depths ranging from 32 to 168 read-pairs enabled unambiguous COI=2 classification for the sample. WGS sequencing coverage, by contrast, detected only moderately elevated SNP heterozygosity (1.9%) in C6-GUY and this elevation was not sufficient to classify COI>1 via The Real McCoil (Chang et al., 2017) (**S7 Fig**.). AMPLseq also identified COI=2 for C6-GUY but without consistent support between replicates (2 vs. 6 biallelic loci). Six additional Guyanese samples were assayed by AMPLseq and one was classified as COI=2. This sample (A5-GUY) gave a stronger minor variant signal in both AMPLseq (15 biallelic loci in both replicates) and WGS data (10.9% SNP heterozygosity) (**S7 Fig**.).

For the Malian sample set, 4CAST and AMPLseq both classified samples E5-PST030 and C6-PST063 as monoclonal and all other samples as polyclonal based on presence/absence of multiallelic loci. While 4CAST detected as many as 10 alternate alleles (median = 3) per sample locus (**Fig. 6B**), AMPLseq detected at most 4 (median = 2). These results reaffirm 4CAST as a tool of choice for resolving higher COI levels and when parasitemia levels are low. AMPLseq may reach simulated performance levels (**S6 Fig**.) by reducing sample multiplexing or sequencing on higher output platforms (*e*.*g*., NovaSeq) for sample sets with low parasitemia.

### Longitudinal sampling: distinguishing recrudescence vs. reinfection

We used 4CAST to examine longitudinal samples that were likely to be diverse and polyclonal. We sequenced samples from the same asymptomatic individual in the longitudinal Mali cohort over three consecutive visits (**Fig. 7**) (Tran et al., 2013). We detected a single microhaplotype at each locus that was present in the first two time points, suggesting a continued infection during the two weeks between samples. At the third time point, we detected a single, distinct microhaplotype at each locus, suggesting that a new infection had occurred and the original infection had disappeared or decreased below our limit of detection (<10 p/µl). In this particular case, the individual was asymptomatic and did not receive anti-malarial treatment between any samples; however, this simple example demonstrates the clarity that 4CAST can bring to tracking infection turnover in longitudinal studies, and suggests its utility in distinguishing recrudescence vs. reinfection in therapeutic efficacy studies.

**Figure 7.**
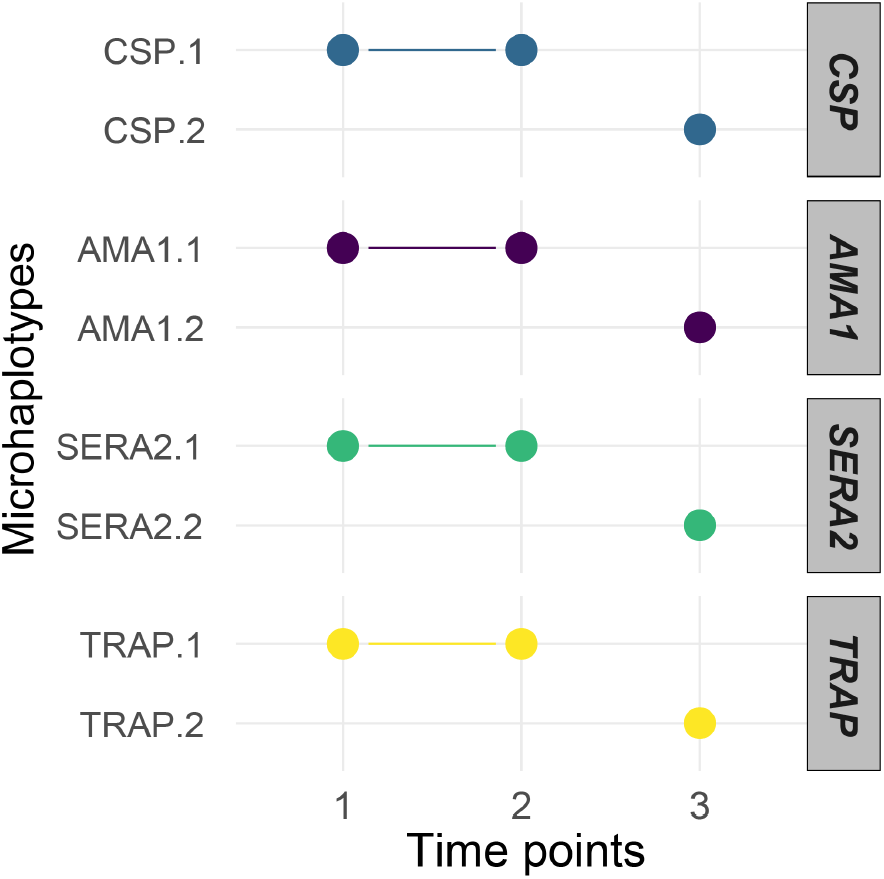
Longitudinal tracking of infections using 4CAST. Identification of distinct microhaplotypes present in samples from an individual at three consecutive time points. The x-axis represents the three time points, and the y-axis represents the individual microhaplotypes identified, grouped by locus. Dots represent the presence of that microhaplotype, connected when present in consecutive visits.

### Drug resistance profiling

AMPLseq loci in *dhfr, mdr1, dhps, kelch13*, and *mdr2* contain ten sequence regions that code for various amino acid (AA) polymorphisms that have previously been associated with resistance to antimalarial drugs (Ariey et al., 2014; Miotto et al., 2015; Mita et al., 2007; Veiga et al., 2016)Thirteen of these 18 positions of interest contained nonsynonymous mutations in Malian and Guyanese clinical samples of this study (Fig. 8). Positions of interest that lacked mutations across both sample sets were DHFR AA 59 and 164; DHPS AA 613; KELCH13 AA 580; and MDR2 AA 484. All Guyanese sequences shared the same mutant alleles at many loci. Malian samples, by contrast, did not show fixed mutant alleles at any amino acid position of interest. A mix of mutant and wildtype alleles occurred among Malian samples for DHFR AA 51 and 108; MDR1 AA 86, 184, and 1246; DHPS AA 436 and 437; and MDR2 AA 492. A previously reported synonymous polymorphism was observed in one Malian sample at KELCH13 *AA* 589 (Taylor et al., 2015).

**Figure 8.**
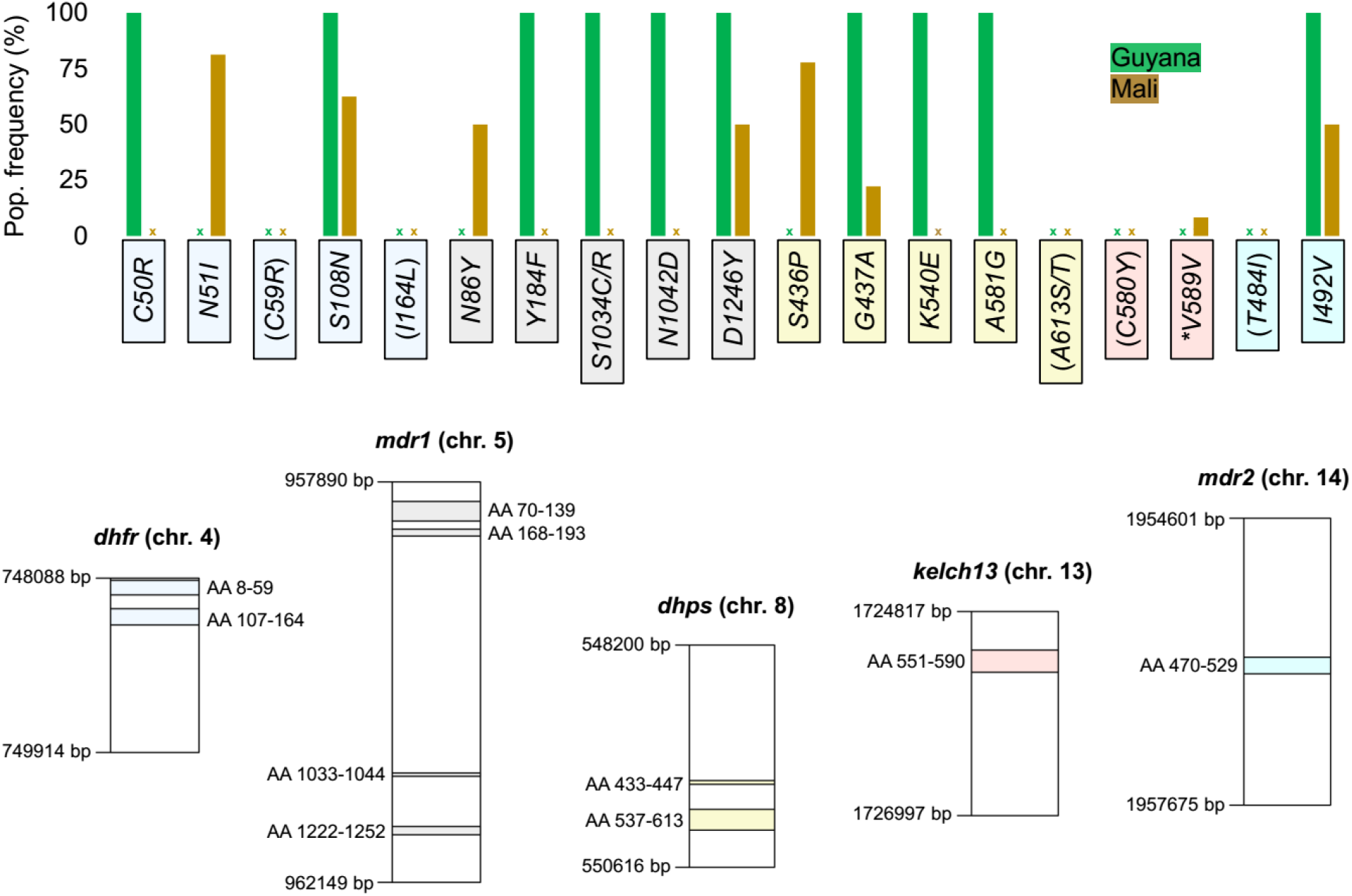
Drug resistance-associated sequence profiling in Guyanese and Malian clinical samples. Bars in the top plot indicate the occurrence of various drug resistance-associated amino acid changes within AMPLseq loci. Positions of interest assayed by AMPLseq but without mutant alleles (see x-marks) in the clinical samples profiled here are labeled in parentheses. Positions 484 and 492 in *mdr2* have been suggested to be involved in artemisinin resistance despite lack of experimental data showing an association with a clinical phenotype (Chenet et al., 2017; Miotto et al., 2015). Bottom plot indicates chromosomal and amino acid (AA) positions of each drug resistance-associated AMPLseq locus. Asterisk indicates synonymous mutation within *kelch13*.

### *P. falciparum and P. vivax* co-infection detection

To test the ability of AMPLseq to detect *P. vivax* co-infections via co-amplification of *PvDHFR*, two additional Guyanese blood spot samples that had been diagnosed as *P. vivax*-only (G4G430) and *P. vivax* + *P. falciparum* co-infection (G4G180) via RDT were included in the sample set. These samples did not undergo sWGA.

*PvDHFR* was detected at high depth in both samples (1068 – 1822 read-pairs for G4G430 and 234 – 560 read-pairs for G4G180) (**Fig. 9**). Only G4G180 also showed read-pair support at *P. falciparum* panel loci (>10 read-pairs at 100 – 115 loci). *PvDHFR* was not detected in any native or pre-amplified 3D7 or mixed-strain (3D7 + Dd2) templates. This demonstrates high specificity of both *PvDHFR* and *P. falciparum* AMPLseq primers to their intended target species without any apparent amplification inhibition by the presence of congeneric DNA.

**Figure 9.**
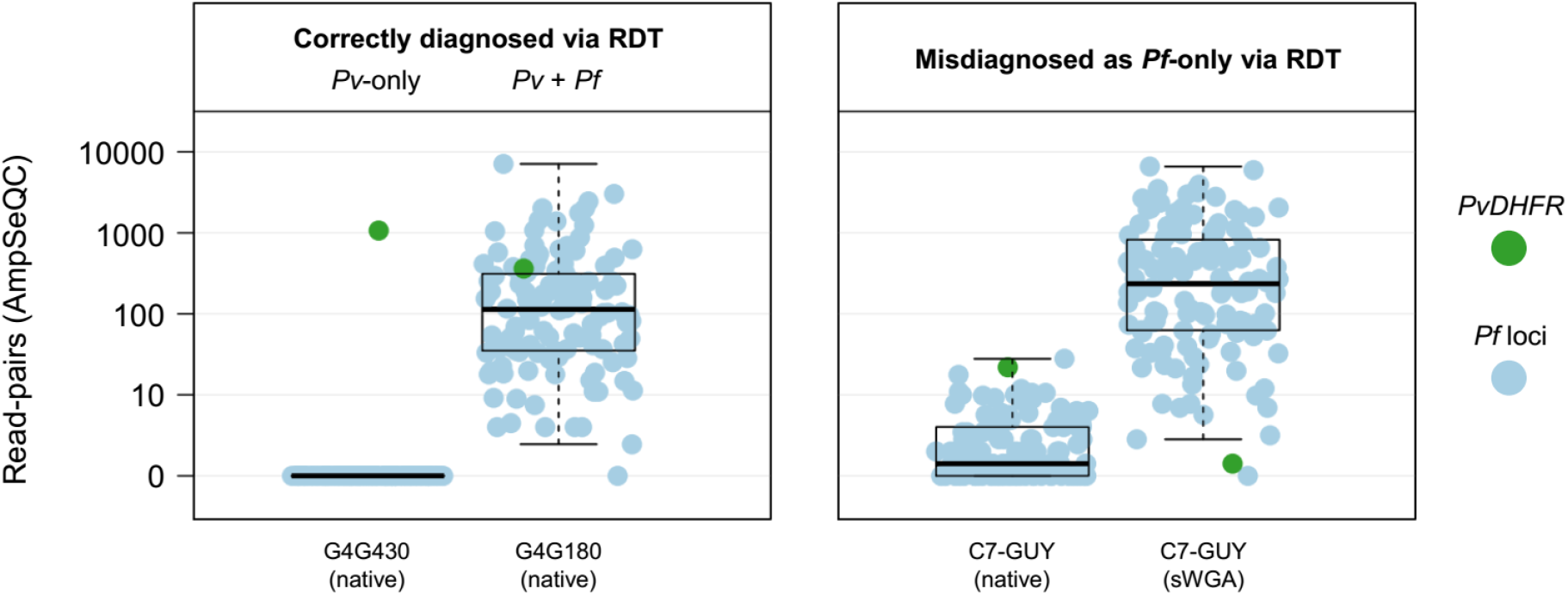
*Plasmodium vivax* detection by AMPLseq. The left panel demonstrates strong read support for *PvDHFR* (green circle) in native control samples previously suggested to contain *P. vivax* (*Pv*; G4G430) and *P. vivax* + *P. falciparum* (*Pv* + *Pf*; G4G180) via RDT. The right panel shows native and sWGA results for C7-GUY, a clinical sample that appears to have been misdiagnosed as *Pf*-only prior to AMPLseq. Blue circles represent read support for *P. falciparum* loci. Positions on the y-axis indicate median read-pair support across two sample replicates. Box and whiskers indicate quartiles.

*PvDHFR* was also detected at low levels (16 – 30 read-pairs) in both native template replicates of C7-GUY, one of the sixteen Guyanese samples previously diagnosed as *P. falciparum*-only via RDT. Surprisingly, two *PvDHFR* read-pairs were also detected in one of the two sWGA replicates from the sample, despite the expectation that sWGA would primarily amplify *P. falciparum* sequencines. Sensitivity of *PvDHFR* detection in pre-amplified samples could be enhanced by adding *PvDHFR* primers to the *P. falciparum* sWGA primer pool. *PvDHFR* detection did not occur in any Malian sample, consistent with low prevalence of *P. vivax in* West Africa relative to the Guiana shield.

## Discussion

The utility of AmpSeq for molecular surveillance of infectious diseases is evidenced by the growing number of protocols recently published or under development for *Plasmodium* and other pathogens (Aydemir et al., 2018; Fola et al., 2020; Jacob et al., 2021; Mitchell et al., 2021; Moser et al., 2021; Ruybal-Pesántez et al., 2021; Schwabl et al., 2020; Tessema et al., 2020). Here, we demonstrate the performance of two new panels for *P. falciparum*, designed to serve different use cases and exhibiting different per-sample costs and levels of complexity. Our comparative analyses of these two new panels, AMPLseq and 4CAST, relative to previously published genotyping panels demonstrates that they perform comparably to existing panels of similar composition across use cases, in a diversity of geographic settings, despite different geographic representation in the population genomic data used to inform their designs. This suggests that *de novo* custom panel design may not be required for accurate COI and relatedness estimation in parasite populations from previously unstudied geographic regions. We therefore suggest that future implementation of these panels should be guided by three criteria: 1) the intended use cases for the data, 2) protocol complexity and compatibility with available instruments and expertise, and 3) protocol customizability for locally relevant genetic loci.

Considering the first of these criteria, intended use case, our investigations above suggest a straightforward mapping of panels by size and feature to use case. The small 4CAST panel is well suited to COI estimation (**Fig. 6**), and profiles four highly diverse antigens for the same effort and cost traditionally used to profile a single locus. Because of the very high diversity of the loci in the 4CAST panel in most parasite populations, this panel is also well suited to any application requiring genetic delineation of distinct parasite lineages (**Fig. 7**). In therapeutic efficacy studies, for example, it is essential to determine whether subjects who become parasitemic following drug treatment are exhibiting a recrudescence of an incompletely-cleared strain from the initial infection (which could indicate treatment failure), or if they have become reinfected with a distinct parasite strain subsequent to treatment. We suggest that the 4CAST panel would be significantly more informative than traditional genotyping approaches used in therapeutic efficacy studies, such as profiling length polymorphisms or allele-specific amplification in the *MSP1*/*MSP2*/*GLURP* loci (Reeder & Marshall, 1994; Snounou, 2002), and more cost-effective than independent monoplex amplification and Illumina sequencing of individual loci (Early et al., 2019; Gruenberg et al., 2019; Lerch et al., 2017).

Our work also demonstrates that the AMPLseq panel performs comparably to two existing multiplexed amplicon sequencing panels of similar size (Jacob et al., 2021; Tessema et al., 2020) for any use case reliant on estimation of parasite relatedness (**Fig. 4**), despite different design criteria and datasets that informed the panels. Potential public health use cases that employ relatedness information include measuring the connectivity of parasites between locations to define units of control, and monitoring changes in the level of transmission (Cerqueira et al., 2017; Daniels et al., 2015; Knudson et al., 2020). The AMPLseq panel and its peers are also much better suited to detecting imported infections given their improved capacity to distinguish parasites from distinct geographic locations (**Fig. 5**.) Finally, the larger panels offer the capacity to monitor genetic markers associated with drug resistance (**Fig. 8**) or, in some panels, detect co-infection with other *Plasmodium* species (**Fig. 9**).

The second panel selection criterion, protocol complexity and compatibility with available instruments, should be prefaced with a reminder that all of these protocols employ nested PCR reactions as the fundamental mechanism to produce sequencing libraries targeting small genomic regions of interest. Any molecular laboratory with a capacity for PCR and a small sequencing instrument such as an Illumina iSeq100 will be capable of carrying out any of these protocols. The exquisite sensitivity of amplicon sequencing implemented via an Illumina platform means that all of these protocols are also susceptible to contamination. PCR reactions should be conducted in dedicated hoods, ideally in rooms or locations physically removed from settings in which PCR products are manipulated. Finally, all of these protocols share: 1) a requirement for careful cleanup of inappropriately large or small DNA molecules from pooled libraries prior to sequencing, 2) precise quantification of said libraries for optimal loading on the sequencing instrument, and 3) large batch sizes (samples in increments of 96, 192, or larger to accommodate the capacity of the intended sequencing instrument).

Though these AmpSeq protocols share many common features, they differ in other aspects that may impact implementation. Whereas the 4CAST and Paragon panels and single nested PCR reactions perform well on native DNA from clinical samples, the AMPLseq and SpotMalaria panels require sWGA pre-amplification prior to the first PCR reaction to ensure adequate performance for samples with parasitemia at or below 100 parasites/µl, which may comprise a significant proportion of samples in some settings. sWGA is an isothermal amplification protocol that is relatively simple to perform but requires an expensive phi29 DNA polymerase and a magnetic bead-based cleanup of individual samples afterward, for an approximate additional cost of $8 USD per sample at the time of writing. Though not large in absolute terms, this cost is comparable to the cost of the AMPLseq or 4CAST protocols themselves, which range from $5 – 10 USD per sample, depending on details of implementation such as sequencing instrument and sample indexing per run. The larger panels additionally employ differing numbers of first-round PCR reactions and require a varying number of magnetic bead-based cleanups to tailor the length profile of intermediate products (summarized in **S5 Table**), which means that the local capacity for automating the bead-based cleanups is a relevant implementation consideration.

The third criterion for panel selection, customizability, may be most relevant for the drug resistance surveillance use case, given differences in the geographic distribution of important drug markers, and varying coverage of known markers by the existing panels. All of the protocols are amenable to customization through the addition of independent target amplifications in the first round of PCR, which could be combined with other first-round PCR multiplex products prior to the second PCR. A more elegant customization approach would be to add (or subtract) targets from the first-round PCR reaction. While complicated bioinformatic pipelines are useful or essential in the design of large multiplexes, in our experience, small multiplexes like 4CAST, which was made from pre-existing primer pairs designed independently, may simply function without optimization, and could presumably be augmented with a small number of additional loci. Though the AMPLseq multiplex of 129 PCR loci benefited from careful design of the original panel, we added the 4CAST targets to the designed AMPLseq target set with no primer modifications and found it to be functional, suggesting it is likely receptive to further augmentation. As the AMPLseq and 4CAST protocols utilize unmodified, commercially available oligos as primers, further customization should be feasible in any setting. However, we must note that not all targets are amenable to incorporation into the multiplex, as we failed despite multiple attempts to include amplicons targeting the *pfcrt* gene associated with chloroquine resistance (Fidock et al., 2000), or the *hrp2/3* genes, which can contain deletions that lead to false-negative diagnosis via rapid diagnostic test (Gamboa et al., 2010).

The proliferation of new AmpSeq protocols for molecular surveillance of infectious diseases raises the important question of whether it is valuable for each disease field to converge on a single approach or common panel. Factors precluding a completely homogeneous approach include varying instrumentation, expertise, and use cases for the data across settings, in addition to an anticipated onward evolution of genotyping technology and elucidation of new markers of interest for drug resistance or other phenotypes. Factors favoring convergence include opportunities for improved procurement of instruments and reagents at a regional level in malaria-endemic countries, and opportunities to directly compare observations between studies and surveillance efforts led by different groups. This latter factor, which we term portability of analyses, has the potential to provide regional or global insight through syntheses across studies. However, the portability of certain analyses is hampered by ascertainment bias, an inherent limitation of any targeted sequencing approach for analyses based on the genotypic state of select loci in different countries. That is, a panel designed based on observations of genetic diversity through WGS in countries A and B may not provide a fair means of comparing diversity in countries C vs. D, if diversity there is distributed differently in the genome than in countries A and B. WGS is the ultimate tool for avoiding this bias. However, the problems of comparing parasite populations profiled with different panels may be mitigated by comparing inferred relatedness levels within populations rather than actual genotypic diversity measures. Overlap of loci among panels would further facilitate direct assessment of relatedness between samples included in different studies (Neafsey, Taylor, & MacInnis, 2021; Taylor et al., 2019). The AMPLseq panel we describe here contains a significant number (n=47) of targets from the SpotMalaria panel, and we expect that future *P. falciparum* panel designs will also tend to exhibit some degree of overlap with other panels, both by deliberate design and through blind convergence based on key genomic features, such as high diversity and sequence amenability to PCR primer design.

As molecular surveillance efforts for malaria and other diseases are more widely adopted and become increasingly diverse, it will be essential for the community to develop standardized approaches for the design, validation, interpretation, and sharing of targeted amplicon sequencing data. The paneljudge R package described here provides an excellent means to comparatively evaluate existing and hypothetical panel performance via data collected from previous population genomic surveys, and the bioinformatic analysis pipelines we have developed are suitable for interpreting Illumina data from diverse targets and panels in different organisms. We anticipate the growth of this field and the development of new analytical tools to extract even more knowledge from increasingly large AmpSeq datasets.

## Data Availability

All amplicon sequencing data, as well as WGS data from 16 Guyana samples, were submitted to the NCBI Sequence Read Archive (http://www.ncbi.nlm.nih.gov/sra) under accession PRJNA758191. This publication uses data from the MalariaGEN Plasmodium falciparum Community Project as described online pending publication and public release of dataset Pf7 (https://www.malariagen.net/resource/34); additional ENA accession numbers are available in Table S4. Previously published data from Guyana and French Guiana can be found at SRA BioProjects PRJNA543530 and PRJNA242182, respectively. Software and documentation can be found at https://github.com/broadinstitute/AmpSeQC (AmpSeQC pipeline), https://github.com/broadinstitute/malaria-amplicon-pipeline.git (Malaria amplicon pipeline), and https://github.com/artaylor85/paneljudge (paneljudge).

https://www.ncbi.nlm.nih.gov/sra/?term=PRJNA758191

https://www.malariagen.net/resource/34

https://www.ncbi.nlm.nih.gov/sra/?term=PRJNA543530

https://www.ncbi.nlm.nih.gov/sra/?term=PRJNA242182

https://github.com/broadinstitute/AmpSeQC

https://github.com/broadinstitute/malaria-amplicon-pipeline.git

https://github.com/artaylor85/paneljudge

## Acknowledgments

This project has been funded in whole or in part with Federal funds from the National Institute of Allergy and Infectious Diseases, National Institutes of Health, Department of Health and Human Services, under Grant Number U19AI110818 to the Broad Institute. This project was also supported by an NIH R01 award to DN (R01AI141544), an award from the Bill and Melinda Gates Foundation to DN and COB (OPP1213366), and a Broad Institute NextGen Award to BM. The Mali cohort study was funded by the Division of Intramural Research, National Institute of Allergy and Infectious Diseases, National Institutes of Health. The Colombian cohort study was supported by British Council Newton-Caldas Fund Institutional Links Award G1854. We thank MalariaGen for use of the Colombian WGS data. We thank Annie Laws for project management. We thank Dr. Nathan Campbell for assistance in the AMPLseq panel design and evaluation.

## Data Accessibility and Benefit Sharing

Data Accessibility: All amplicon sequencing data, as well as WGS data from 16 Guyana samples, were submitted to the NCBI Sequence Read Archive (http://www.ncbi.nlm.nih.gov/sra) under accession PRJNA758191. This publication uses data from the MalariaGEN Plasmodium falciparum Community Project as described online pending publication and public release of dataset Pf7 (https://www.malariagen.net/resource/34); additional ENA accession numbers are available in Table S4. Previously published data from Guyana and French Guiana can be found at SRA BioProjects PRJNA543530 and PRJNA242182, respectively. Software and documentation can be found at https://github.com/broadinstitute/AmpSeQC (AmpSeQC pipeline), https://github.com/broadinstitute/malaria-amplicon-pipeline.git (Malaria amplicon pipeline), and https://github.com/artaylor85/paneljudge (paneljudge).

Benefits Generated: A research collaboration was developed with scientists from the countries providing genetic samples, all collaborators are included as co-authors, the results of research have been shared with the provider communities and the broader scientific community (see above), and the research addresses a priority concern, in this case the public health concern of malaria. More broadly, our group is committed to international scientific partnerships, as well as institutional capacity building.

## Author Contributions

- Conceptualization EL, PS, MC, ART, AME, BLM, DEN
- Data Curation ZMJ, RP, TJS
- Formal Analysis EL, PS, MC, ART, RP, TJS
- Funding Acquisition COB, BLM, DEN
- Investigation PS, MC, ZMJ, MS, RK, SW
- Methodology EL, MC, PS, ART, ZMJ, MS, RP, TJS, RK
- Project Administration BLM, DEN
- Resources CMA, SP, PDC, BT, JCR, VC, KJ, HC
- Software ART, RP, TJS, AME
- Supervision COB, BLM, AME, DEN
- Validation EL, PS, MC, ZMJ, MS, RP, TJS, RK
- Visualization EL, PS, MC, ART, RP
- Writing - Original Draft Preparation EL, PS, MC, ART, DEN
- Writing - Review & Editing [All authors]

## References

Ariey, F., Witkowski, B., Amaratunga, C., Beghain, J., Langlois, A.-C., Khim, N., Kim, S., Duru, V., Bouchier, C., Ma, L., Lim, P., Leang, R., Duong, S., Sreng, S., Suon, S., Chuor, C. M., Bout, D. M., Ménard, S., Rogers, W. O., … Ménard, D. (2014). A molecular marker of artemisinin-resistant Plasmodium falciparum malaria. Nature, 505(7481), 50–55. https://doi.org/10.1038/nature12876

Aydemir, O., Janko, M., Hathaway, N. J., Verity, R., Mwandagalirwa, M. K., Tshefu, A. K., Tessema, S. K., Marsh, P. W., Tran, A., Reimonn, T., Ghani, A. C., Ghansah, A., Juliano, J. J., Greenhouse, B. R., Emch, M., Meshnick, S. R., & Bailey, J. A. (2018). Drug-Resistance and Population Structure of Plasmodium falciparum Across the Democratic Republic of Congo Using High-Throughput Molecular Inversion Probes. The Journal of Infectious Diseases, 218(6), 946–955. https://doi.org/10.1093/infdis/jiy223

Baetscher, D. S., Clemento, A. J., Ng, T. C., Anderson, E. C., & Garza, J. C. (2018). Microhaplotypes provide increased power from short-read DNA sequences for relationship inference. Molecular Ecology Resources, 18(2), 296–305. https://doi.org/10.1111/1755-0998.12737

Baker, S., Thomson, N., Weill, F.-X., & Holt, K. E. (2018). Genomic insights into the emergence and spread of antimicrobial-resistant bacterial pathogens. Science, 360(6390), 733–738. https://doi.org/10.1126/science.aar3777

Baniecki, M. L., Faust, A. L., Schaffner, S. F., Park, D. J., Galinsky, K., Daniels, R. F., Hamilton, E., Ferreira, M. U., Karunaweera, N. D., Serre, D., Zimmerman, P. A., Sá, J. M., Wellems, T. E., Musset, L., Legrand, E., Melnikov, A., Neafsey, D. E., Volkman, S. K., Wirth, D. F., & Sabeti, P. C. (2015). Development of a single nucleotide polymorphism barcode to genotype Plasmodium vivax infections. PLoS Neglected Tropical Diseases, 9(3), e0003539. https://doi.org/10.1371/journal.pntd.0003539

Callahan, B. J., McMurdie, P. J., Rosen, M. J., Han, A. W., Johnson, A. J. A., & Holmes, S. P. (2016). DADA2: High-resolution sample inference from Illumina amplicon data. Nature Methods, 13(7), 581–583. https://doi.org/10.1038/nmeth.3869

Campbell, N. R., Harmon, S. A., & Narum, S. R. (2015). Genotyping-in-Thousands by sequencing (GT-seq): A cost effective SNP genotyping method based on custom amplicon sequencing. Molecular Ecology Resources, 15(4), 855–867. https://doi.org/10.1111/1755-0998.12357

Cerqueira, G. C., Cheeseman, I. H., Schaffner, S. F., Nair, S., McDew-White, M., Phyo, A. P., Ashley, E. A., Melnikov, A., Rogov, P., Birren, B. W., Nosten, F., Anderson, T. J. C., & Neafsey, D. E. (2017). Longitudinal genomic surveillance of Plasmodium falciparum malaria parasites reveals complex genomic architecture of emerging artemisinin resistance. Genome Biology, 18(1), 78. https://doi.org/10.1186/s13059-017-1204-4

Chang, H.-H., Wesolowski, A., Sinha, I., Jacob, C. G., Mahmud, A., Uddin, D., Zaman, S. I., Hossain, M. A., Faiz, M. A., Ghose, A., Sayeed, A. A., Rahman, M. R., Islam, A., Karim, M. J., Rezwan, M. K., Shamsuzzaman, A. K. M., Jhora, S. T., Aktaruzzaman, M. M., Drury, E., … Buckee, C. (2019). Mapping imported malaria in Bangladesh using parasite genetic and human mobility data. ELife, 8, e43481. https://doi.org/10.7554/eLife.43481

Chang, H.-H., Worby, C. J., Yeka, A., Nankabirwa, J., Kamya, M. R., Staedke, S. G., Dorsey, G., Murphy, M., Neafsey, D. E., Jeffreys, A. E., Hubbart, C., Rockett, K. A., Amato, R., Kwiatkowski, D. P., Buckee, C. O., & Greenhouse, B. (2017). THE REAL McCOIL: A method for the concurrent estimation of the complexity of infection and SNP allele frequency for malaria parasites. PLoS Computational Biology, 13(1), e1005348. https://doi.org/10.1371/journal.pcbi.1005348

Chenet, S. M., Akinyi Okoth, S., Huber, C. S., Chandrabose, J., Lucchi, N. W., Talundzic, E., Krishnalall, K., Ceron, N., Musset, L., Macedo de Oliveira, A., Venkatesan, M., Rahman, R., Barnwell, J. W., & Udhayakumar, V. (2016). Independent Emergence of the Plasmodium falciparum Kelch Propeller Domain Mutant Allele C580Y in Guyana. The Journal of Infectious Diseases, 213(9), 1472–1475. https://doi.org/10.1093/infdis/jiv752

Chenet, S. M., Okoth, S. A., Kelley, J., Lucchi, N., Huber, C. S., Vreden, S., Macedo de Oliveira, A., Barnwell, J. W., Udhayakumar, V., & Adhin, M. R. (2017). Molecular Profile of Malaria Drug Resistance Markers of Plasmodium falciparum in Suriname. Antimicrobial Agents and Chemotherapy, 61(7), e02655–16. https://doi.org/10.1128/AAC.02655-16

Coll, F., Harrison, E. M., Toleman, M. S., Reuter, S., Raven, K. E., Blane, B., Palmer, B., Kappeler, A. R. M., Brown, N. M., Török, M. E., Parkhill, J., & Peacock, S. J. (2017). Longitudinal genomic surveillance of MRSA in the UK reveals transmission patterns in hospitals and the community. Science Translational Medicine, 9(413), eaak9745. https://doi.org/10.1126/scitranslmed.aak9745

Dalmat, R., Naughton, B., Kwan-Gett, T. S., Slyker, J., & Stuckey, E. M. (2019). Use cases for genetic epidemiology in malaria elimination. Malaria Journal, 18(1), 1–11. https://doi.org/10.1186/s12936-019-2784-0

Danecek, P., Auton, A., Abecasis, G., Albers, C. A., Banks, E., DePristo, M. A., Handsaker, R. E., Lunter, G., Marth, G. T., Sherry, S. T., McVean, G., Durbin, R., & 1000 Genomes Project Analysis Group. (2011). The variant call format and VCFtools. Bioinformatics, 27(15), 2156–2158. https://doi.org/10.1093/bioinformatics/btr330

Daniels, R. F., Schaffner, S. F., Wenger, E. A., Proctor, J. L., Chang, H.-H., Wong, W., Baro, N., Ndiaye, D., Fall, F. B., Ndiop, M., Ba, M., Milner, D. A., Taylor, T. E., Neafsey, D. E., Volkman, S. K., Eckhoff, P. A., Hartl, D. L., & Wirth, D. F. (2015). Modeling malaria genomics reveals transmission decline and rebound in Senegal. Proceedings of the National Academy of Sciences of the United States of America, 112(22), 7067–7072. https://doi.org/10.1073/pnas.1505691112

Daniels, R., Volkman, S. K., Milner, D. A., Mahesh, N., Neafsey, D. E., Park, D. J., Rosen, D., Angelino, E., Sabeti, P. C., Wirth, D. F., & Wiegand, R. C. (2008). A general SNP-based molecular barcode for Plasmodium falciparum identification and tracking. Malaria Journal, 7, 223. https://doi.org/10.1186/1475-2875-7-223

DePristo, M. A., Banks, E., Poplin, R., Garimella, K. V., Maguire, J. R., Hartl, C., Philippakis, A. A., del Angel, G., Rivas, M. A., Hanna, M., McKenna, A., Fennell, T. J., Kernytsky, A. M., Sivachenko, A. Y., Cibulskis, K., Gabriel, S. B., Altshuler, D., & Daly, M. J. (2011). A framework for variation discovery and genotyping using next-generation DNA sequencing data. Nature Genetics, 43(5), 491–498. https://doi.org/10.1038/ng.806

Early, A. M., Daniels, R. F., Farrell, T. M., Grimsby, J., Volkman, S. K., Wirth, D. F., MacInnis, B. L., & Neafsey, D. E. (2019). Detection of low-density Plasmodium falciparum infections using amplicon deep sequencing. Malaria Journal, 18(1), 219. https://doi.org/10.1186/s12936-019-2856-1

Fidock, D. A., Nomura, T., Talley, A. K., Cooper, R. A., Dzekunov, S. M., Ferdig, M. T., Ursos, L. M., Sidhu, A. B., Naudé, B., Deitsch, K. W., Su, X. Z., Wootton, J. C., Roepe, P. D., & Wellems, T. E. (2000). Mutations in the P. falciparum digestive vacuole transmembrane protein PfCRT and evidence for their role in chloroquine resistance. Molecular Cell, 6(4), 861–871. https://doi.org/10.1016/s1097-2765(05)00077-8

Fola, A. A., Kattenberg, E., Razook, Z., Lautu-Gumal, D., Lee, S., Mehra, S., Bahlo, M., Kazura, J., Robinson, L. J., Laman, M., Mueller, I., & Barry, A. E. (2020). SNP barcodes provide higher resolution than microsatellite markers to measure Plasmodium vivax population genetics. Malaria Journal, 19(1), 375. https://doi.org/10.1186/s12936-020-03440-0

Galinsky, K., Valim, C., Salmier, A., de Thoisy, B., Musset, L., Legrand, E., Faust, A., Baniecki, M., Ndiaye, D., Daniels, R. F., Hartl, D. L., Sabeti, P. C., Wirth, D. F., Volkman, S. K., & Neafsey, D. E. (2015). COIL: a methodology for evaluating malarial complexity of infection using likelihood from single nucleotide polymorphism data. Malaria Journal, 14(1), 4. https://doi.org/10.1186/1475-2875-14-4

Gamboa, D., Ho, M.-F., Bendezu, J., Torres, K., Chiodini, P. L., Barnwell, J. W., Incardona, S., Perkins, M., Bell, D., McCarthy, J., & Cheng, Q. (2010). A Large Proportion of P. falciparum Isolates in the Amazon Region of Peru Lack pfhrp2 and pfhrp3: Implications for Malaria Rapid Diagnostic Tests. PloS One, 5(1). https://doi.org/10.1371/journal.pone.0008091

Gruenberg, M., Lerch, A., Beck, H.-P., & Felger, I. (2019). Amplicon deep sequencing improves Plasmodium falciparum genotyping in clinical trials of antimalarial drugs. Scientific Reports, 9(1), 1–12. https://doi.org/10.1038/s41598-019-54203-0

Hargrove, J. S., McCane, J., Roth, C. J., High, B., & Campbell, M. R. (2021). Mating systems and predictors of relative reproductive success in a Cutthroat Trout subspecies of conservation concern. Ecology and Evolution, 11(16). https://doi.org/10.1002/ece3.7914

Helb, D. A., Tetteh, K. K. A., Felgner, P. L., Skinner, J., Hubbard, A., Arinaitwe, E., Mayanja-Kizza, H., Ssewanyana, I., Kamya, M. R., Beeson, J. G., Tappero, J., Smith, D. L., Crompton, P. D., Rosenthal, P. J., Dorsey, G., Drakeley, C. J., & Greenhouse, B. (2015). Novel serologic biomarkers provide accurate estimates of recent Plasmodium falciparum exposure for individuals and communities. Proceedings of the National Academy of Sciences, 112(32), E4438–E4447. https://doi.org/10.1073/pnas.1501705112

Henden, L., Lee, S., Mueller, I., Barry, A., & Bahlo, M. (2018). Identity-by-descent analyses for measuring population dynamics and selection in recombining pathogens. PLoS Genetics, 14(5), e1007279. https://doi.org/10.1371/journal.pgen.1007279

Jacob, C. G., Thuy-Nhien, N., Mayxay, M., Maude, R. J., Quang, H. H., Hongvanthong, B., Vanisaveth, V., Ngo Duc, T., Rekol, H., van der Pluijm, R., von Seidlein, L., Fairhurst, R., Nosten, F., Hossain, M. A., Park, N., Goodwin, S., Ringwald, P., Chindavongsa, K., Newton, P., … Miotto, O. (2021). Genetic surveillance in the Greater Mekong subregion and South Asia to support malaria control and elimination. ELife, 10, e62997. https://doi.org/10.7554/eLife.62997

Jones, S., Kay, K., Hodel, E. M., Gruenberg, M., Lerch, A., Felger, I., & Hastings, I. (2021). Should deep-sequenced amplicons become the new gold-standard for analysing malaria drug clinical trials? Antimicrobial Agents and Chemotherapy, AAC0043721. https://doi.org/10.1128/AAC.00437-21

Kayiba, N. K., Yobi, D. M., Tshibangu-Kabamba, E., Tuan, V. P., Yamaoka, Y., Devleesschauwer, B., Mvumbi, D. M., Okitolonda Wemakoy, E., De Mol, P., Mvumbi, G. L., Hayette, M.-P., Rosas-Aguirre, A., & Speybroeck, N. (2021). Spatial and molecular mapping of Pfkelch13 gene polymorphism in Africa in the era of emerging Plasmodium falciparum resistance to artemisinin: a systematic review. The Lancet. Infectious Diseases, 21(4), e82–e92. https://doi.org/10.1016/S1473-3099(20)30493-X

Knudson, A., González-Casabianca, F., Feged-Rivadeneira, A., Pedreros, M. F., Aponte, S., Olaya, A., Castillo, C. F., Mancilla, E., Piamba-Dorado, A., Sanchez-Pedraza, R., Salazar-Terreros, M. J., Lucchi, N., Udhayakumar, V., Jacob, C., Pance, A., Carrasquilla, M., Apráez, G., Angel, J. A., Rayner, J. C., & Corredor, V. (2020). Spatio-temporal dynamics of Plasmodium falciparum transmission within a spatial unit on the Colombian Pacific Coast. Scientific Reports, 10(1), 3756. https://doi.org/10.1038/s41598-020-60676-1

Krijthe, J. H. (2015). Rtsne: T-Distributed Stochastic Neighbor Embedding using a Barnes-Hut Implementation. https://github.com/jkrijthe/Rtsne

Lautu-Gumal, D., Razook, Z., Koleala, T., Nate, E., McEwen, S., Timbi, D., Hetzel, M. W., Lavu, E., Tefuarani, N., Makita, L., Kazura, J., Mueller, I., Pomat, W., Laman, M., Robinson, L. J., & Barry, A. E. (2021). Surveillance of molecular markers of Plasmodium falciparum artemisinin resistance (kelch13 mutations) in Papua New Guinea between 2016 and 2018. International Journal for Parasitology. Drugs and Drug Resistance, 16, 188–193. https://doi.org/10.1016/j.ijpddr.2021.06.004

Lefterova, M. I., Budvytiene, I., Sandlund, J., Färnert, A., & Banaei, N. (2015). Simple real-time PCR and amplicon sequencing method for identification of plasmodium species in human whole blood. Journal of Clinical Microbiology, 53(7), 2251–2257. https://doi.org/10.1128/JCM.00542-15

Lerch, A., Koepfli, C., Hofmann, N. E., Messerli, C., Wilcox, S., Kattenberg, J. H., Betuela, I., O’Connor, L., Mueller, I., & Felger, I. (2017). Development of amplicon deep sequencing markers and data analysis pipeline for genotyping multi-clonal malaria infections. BMC Genomics, 18(1), 864. https://doi.org/10.1186/s12864-017-4260-y

Li, H. (2013). Aligning sequence reads, clone sequences and assembly contigs with BWA-MEM. ArXiv:1303.3997 [Preprint]. http://arxiv.org/abs/1303.3997

Liu, Y., Tessema, S. K., Murphy, M., Xu, S., Schwartz, A., Wang, W., Cao, Y., Lu, F., Tang, J., Gu, Y., Zhu, G., Zhou, H., Gao, Q., Huang, R., Cao, J., & Greenhouse, B. (2020). Confirmation of the absence of local transmission and geographic assignment of imported falciparum malaria cases to China using microsatellite panel. Malaria Journal, 19(1), 244. https://doi.org/10.1186/s12936-020-03316-3

MalariaGEN Plasmodium falciparum Community Project. (2016). Genomic epidemiology of artemisinin resistant malaria. ELife, 5, e08714. https://doi.org/10.7554/eLife.08714

Mathieu, L. C., Cox, H., Early, A. M., Mok, S., Lazrek, Y., Paquet, J.-C., Ade, M.-P., Lucchi, N. W., Grant, Q., Udhayakumar, V., Alexandre, J. S., Demar, M., Ringwald, P., Neafsey, D. E., Fidock, D. A., & Musset, L. (2020). Local emergence in Amazonia of Plasmodium falciparum k13 C580Y mutants associated with in vitro artemisinin resistance. ELife, 9, e51015. https://doi.org/10.7554/eLife.51015

McKenna, A., Hanna, M., Banks, E., Sivachenko, A., Cibulskis, K., Kernytsky, A., Garimella, K., Altshuler, D., Gabriel, S., Daly, M., & DePristo, M. A. (2010). The Genome Analysis Toolkit: a MapReduce framework for analyzing next-generation DNA sequencing data. Genome Research, 20(9), 1297–1303. https://doi.org/10.1101/gr.107524.110

Miles, A., Iqbal, Z., Vauterin, P., Pearson, R., Campino, S., Theron, M., Gould, K., Mead, D., Drury, E., O’Brien, J., Rubio, V. R., MacInnis, B., Mwangi, J., Samarakoon, U., Ranford-Cartwright, L., Ferdig, M., Hayton, K., Su, X., Wellems, T., … Kwiatkowski, D. (2016). Indels, structural variation, and recombination drive genomic diversity in Plasmodium falciparum. Genome Research, 26(9), 1288–1299. https://doi.org/10.1101/gr.203711.115

Miles, A., pyup.io bot, Murillo, R., Ralph, P., Harding, N. J., Rahul Pisupati, Summer Rae, & Tim Millar. (2020). cggh/scikit-allel: v1.3.2. Zenodo. https://doi.org/10.5281/zenodo.3976233

Miller, R. H., Hathaway, N. J., Kharabora, O., Mwandagalirwa, K., Tshefu, A., Meshnick, S. R., Taylor, S. M., Juliano, J. J., Stewart, V. A., & Bailey, J. A. (2017). A deep sequencing approach to estimate Plasmodium falciparum complexity of infection (COI) and explore apical membrane antigen 1 diversity. Malaria Journal, 16(1), 490. https://doi.org/10.1186/s12936-017-2137-9

Miotto, O., Amato, R., Ashley, E. A., MacInnis, B., Almagro-Garcia, J., Amaratunga, C., Lim, P., Mead, D., Oyola, S. O., Dhorda, M., Imwong, M., Woodrow, C., Manske, M., Stalker, J., Drury, E., Campino, S., Amenga-Etego, L., Thanh, T.-N. N., Tran, H. T., … Kwiatkowski, D. P. (2015). Genetic architecture of artemisinin-resistant Plasmodium falciparum. Nature Genetics, 47(3), 226–234. https://doi.org/10.1038/ng.3189

Miotto, O., Sekihara, M., Tachibana, S.-I., Yamauchi, M., Pearson, R. D., Amato, R., Gonçalves, S., Mehra, S., Noviyanti, R., Marfurt, J., Auburn, S., Price, R. N., Mueller, I., Ikeda, M., Mori, T., Hirai, M., Tavul, L., Hetzel, M. W., Laman, M., … Mita, T. (2020). Emergence of artemisinin-resistant Plasmodium falciparum with kelch13 C580Y mutations on the island of New Guinea. PLoS Pathogens, 16(12), e1009133. https://doi.org/10.1371/journal.ppat.1009133

Mita, T., Tanabe, K., Takahashi, N., Tsukahara, T., Eto, H., Dysoley, L., Ohmae, H., Kita, K., Krudsood, S., Looareesuwan, S., Kaneko, A., Björkman, A., & Kobayakawa, T. (2007). Independent Evolution of Pyrimethamine Resistance in Plasmodium falciparum Isolates in Melanesia. Antimicrobial Agents and Chemotherapy, 51(3), 1071–1077. https://doi.org/10.1128/AAC.01186-06

Mitchell, R. M., Zhou, Z., Sheth, M., Sergent, S., Frace, M., Nayak, V., Hu, B., Gimnig, J., ter Kuile, F., Lindblade, K., Slutsker, L., Hamel, M. J., Desai, M., Otieno, K., Kariuki, S., Vigfusson, Y., & Shi, Y. P. (2021). Development of a new barcode-based, multiplex-PCR, next-generation-sequencing assay and data processing and analytical pipeline for multiplicity of infection detection of Plasmodium falciparum. Malaria Journal, 20(1), 92. https://doi.org/10.1186/s12936-021-03624-2

Moser, K. A., Madebe, R. A., Aydemir, O., Chiduo, M. G., Mandara, C. I., Rumisha, S. F., Chaky, F., Denton, M., Marsh, P. W., Verity, R., Watson, O. J., Ngasala, B., Mkude, S., Molteni, F., Njau, R., Warsame, M., Mandike, R., Kabanywanyi, A. M., Mahende, M. K., … Bailey, J. A. (2021). Describing the current status of Plasmodium falciparum population structure and drug resistance within mainland Tanzania using molecular inversion probes. Molecular Ecology, 30(1), 100–113. https://doi.org/10.1111/mec.15706

Natesh, M., Taylor, R. W., Truelove, N. K., Hadly, E. A., Palumbi, S. R., Petrov, D. A., & Ramakrishnan, U. (2019). Empowering conservation practice with efficient and economical genotyping from poor quality samples. Methods in Ecology and Evolution, 10(6), 853–859. https://doi.org/10.1111/2041-210X.13173

Neafsey, D. E., Juraska, M., Bedford, T., Benkeser, D., Valim, C., Griggs, A., Lievens, M., Abdulla, S., Adjei, S., Agbenyega, T., Agnandji, S. T., Aide, P., Anderson, S., Ansong, D., Aponte, J. J., Asante, K. P., Bejon, P., Birkett, A. J., Bruls, M., … Wirth, D. F. (2015). Genetic Diversity and Protective Efficacy of the RTS,S/AS01 Malaria Vaccine. New England Journal of Medicine, 373(21), 2025–2037. https://doi.org/10.1056/NEJMoa1505819

Neafsey, D. E., Taylor, A. R., & MacInnis, B. L. (2021). Advances and opportunities in malaria population genomics. Nature Reviews. Genetics. https://doi.org/10.1038/s41576-021-00349-5

Nelson, C. S., Sumner, K. M., Freedman, E., Saelens, J. W., Obala, A. A., Mangeni, J. N., Taylor, S. M., & O’Meara, W. P. (2019). High-resolution micro-epidemiology of parasite spatial and temporal dynamics in a high malaria transmission setting in Kenya. Nature Communications, 10, 5615. https://doi.org/10.1038/s41467-019-13578-4

Oude Munnink, B. B., Worp, N., Nieuwenhuijse, D. F., Sikkema, R. S., Haagmans, B., Fouchier, R. A. M., & Koopmans, M. (2021). The next phase of SARS-CoV-2 surveillance: real-time molecular epidemiology. Nature Medicine. https://doi.org/10.1038/s41591-021-01472-w

Oyola, S. O., Ariani, C. V., Hamilton, W. L., Kekre, M., Amenga-Etego, L. N., Ghansah, A., Rutledge, G. G., Redmond, S., Manske, M., Jyothi, D., Jacob, C. G., Otto, T. D., Rockett, K., Newbold, C. I., Berriman, M., & Kwiatkowski, D. P. (2016). Whole genome sequencing of Plasmodium falciparum from dried blood spots using selective whole genome amplification. Malaria Journal, 15(1). https://doi.org/10.1186/s12936-016-1641-7

Pelleau, S., Moss, E. L., Dhingra, S. K., Volney, B., Casteras, J., Gabryszewski, S. J., Volkman, S. K., Wirth, D. F., Legrand, E., Fidock, D. A., Neafsey, D. E., & Musset, L. (2015). Adaptive evolution of malaria parasites in French Guiana: Reversal of chloroquine resistance by acquisition of a mutation in pfcrt. Proceedings of the National Academy of Sciences, 112(37), 11672–11677. https://doi.org/10.1073/pnas.1507142112

Quick, J., Loman, N. J., Duraffour, S., Simpson, J. T., Severi, E., Cowley, L., Bore, J. A., Koundouno, R., Dudas, G., Mikhail, A., Ouédraogo, N., Afrough, B., Bah, A., Baum, J. H. J., Becker-Ziaja, B., Boettcher, J. P., Cabeza-Cabrerizo, M., Camino-Sánchez, Á., Carter, L. L., … Carroll, M. W. (2016). Real-time, portable genome sequencing for Ebola surveillance. Nature, 530(7589), 228–232. https://doi.org/10.1038/nature16996

Reeder, J. C., & Marshall, V. M. (1994). A simple method for typing Plasmodium falciparum merozoite surface antigens 1 and 2 (MSA-1 and MSA-2) using a dimorphic-form specific polymerase chain reaction. Molecular and Biochemical Parasitology, 68(2), 329–332. https://doi.org/10.1016/0166-6851(94)90179-1

Ruybal-Pesántez, S., Sáenz, F. E., Deed, S., Johnson, E. K., Larremore, D. B., Vera-Arias, C. A., Tiedje, K. E., & Day, K. P. (2021). Clinical malaria incidence following an outbreak in Ecuador was predominantly associated with Plasmodium falciparum with recombinant variant antigen gene repertoires. MedRxiv [Preprint], 2021.04.12.21255093. https://doi.org/10.1101/2021.04.12.21255093

Schaffner, S. F., Taylor, A. R., Wong, W., Wirth, D. F., & Neafsey, D. E. (2018). hmmIBD: software to infer pairwise identity by descent between haploid genotypes. Malaria Journal, 17(1), 196. https://doi.org/10.1186/s12936-018-2349-7

Schmidt, D. A., Campbell, N. R., Govindarajulu, P., Larsen, K. W., & Russello, M. A. (2020). Genotyping-in-Thousands by sequencing (GT-seq) panel development and application to minimally invasive DNA samples to support studies in molecular ecology. Molecular Ecology Resources, 20(1), 114–124. https://doi.org/10.1111/1755-0998.13090

Schwabl, P., Maiguashca Sánchez, J., Costales, J. A., Ocaña-Mayorga, S., Segovia, M., Carrasco, H. J., Hernández, C., Ramírez, J. D., Lewis, M. D., Grijalva, M. J., & Llewellyn, M. S. (2020). Culture-free genome-wide locus sequence typing (GLST) provides new perspectives on Trypanosoma cruzi dispersal and infection complexity. PLoS Genetics, 16(12), e1009170. https://doi.org/10.1371/journal.pgen.1009170

Snounou, G. (2002). Genotyping of Plasmodium spp. Nested PCR. Methods in Molecular Medicine, 72, 103–116. https://doi.org/10.1385/1-59259-271-6:103

Takala-Harrison, S., Jacob, C. G., Arze, C., Cummings, M. P., Silva, J. C., Dondorp, A. M., Fukuda, M. M., Hien, T. T., Mayxay, M., Noedl, H., Nosten, F., Kyaw, M. P., Nhien, N. T. T., Imwong, M., Bethell, D., Se, Y., Lon, C., Tyner, S. D., Saunders, D. L., … Plowe, C. V. (2015). Independent emergence of artemisinin resistance mutations among Plasmodium falciparum in Southeast Asia. The Journal of Infectious Diseases, 211(5), 670–679. https://doi.org/10.1093/infdis/jiu491

Taylor, A. R., & Jacob, P. E. (2020). paneljudge: Judge the performance of a panel of genetic markers using simulated data. (R package version 0.0.0.9000) [Computer software].

Taylor, A. R., Jacob, P. E., Neafsey, D. E., & Buckee, C. O. (2019). Estimating Relatedness Between Malaria Parasites. Genetics, 212(4), 1337–1351. https://doi.org/10.1534/genetics.119.302120

Taylor, A. R., Schaffner, S. F., Cerqueira, G. C., Nkhoma, S. C., Anderson, T. J. C., Sriprawat, K., Pyae Phyo, A., Nosten, F., Neafsey, D. E., & Buckee, C. O. (2017). Quantifying connectivity between local Plasmodium falciparum malaria parasite populations using identity by descent. PLoS Genetics, 13(10), e1007065. https://doi.org/10.1371/journal.pgen.1007065

Taylor, S. M., Parobek, C. M., DeConti, D. K., Kayentao, K., Coulibaly, S. O., Greenwood, B. M., Tagbor, H., Williams, J., Bojang, K., Njie, F., Desai, M., Kariuki, S., Gutman, J., Mathanga, D. P., Mårtensson, A., Ngasala, B., Conrad, M. D., Rosenthal, P. J., Tshefu, A. K., … Juliano, J. J. (2015). Absence of Putative Artemisinin Resistance Mutations Among Plasmodium falciparum in Sub-Saharan Africa: A Molecular Epidemiologic Study. The Journal of Infectious Diseases, 211(5), 680–688. https://doi.org/10.1093/infdis/jiu467

Tessema, S. K., Hathaway, N. J., Teyssier, N. B., Murphy, M., Chen, A., Aydemir, O., Duarte, E. M., Simone, W., Colborn, J., Saute, F., Crawford, E., Aide, P., Bailey, J. A., & Greenhouse, B. (2020). Sensitive, highly multiplexed sequencing of microhaplotypes from the Plasmodium falciparum heterozygome. The Journal of Infectious Diseases. https://doi.org/10.1093/infdis/jiaa527

Tessema, S., Wesolowski, A., Chen, A., Murphy, M., Wilheim, J., Mupiri, A.-R., Ruktanonchai, N. W., Alegana, V. A., Tatem, A. J., Tambo, M., Didier, B., Cohen, J. M., Bennett, A., Sturrock, H. J., Gosling, R., Hsiang, M. S., Smith, D. L., Mumbengegwi, D. R., Smith, J. L., & Greenhouse, B. (2019). Using parasite genetic and human mobility data to infer local and cross-border malaria connectivity in Southern Africa. ELife, 8, e43510. https://doi.org/10.7554/eLife.43510

Trager, W., & Jensen, J. B. (1976). Human malaria parasites in continuous culture. Science (New York, N.Y.), 193(4254), 673–675. https://doi.org/10.1126/science.781840

Tran, T. M., Li, S., Doumbo, S., Doumtabe, D., Huang, C.-Y., Dia, S., Bathily, A., Sangala, J., Kone, Y., Traore, A., Niangaly, M., Dara, C., Kayentao, K., Ongoiba, A., Doumbo, O. K., Traore, B., & Crompton, P. D. (2013). An Intensive Longitudinal Cohort Study of Malian Children and Adults Reveals No Evidence of Acquired Immunity to Plasmodium falciparum Infection. Clinical Infectious Diseases: An Official Publication of the Infectious Diseases Society of America, 57(1), 40–47. https://doi.org/10.1093/cid/cit174

Van der Auwera, G. A., Carneiro, M. O., Hartl, C., Poplin, R., Del Angel, G., Levy-Moonshine, A., Jordan, T., Shakir, K., Roazen, D., Thibault, J., Banks, E., Garimella, K. V., Altshuler, D., Gabriel, S., & DePristo, M. A. (2013). From FastQ data to high confidence variant calls: the Genome Analysis Toolkit best practices pipeline. Current Protocols in Bioinformatics, 43, 11.10.1-11.10.33. https://doi.org/10.1002/0471250953.bi1110s43

Veiga, M. I., Dhingra, S. K., Henrich, P. P., Straimer, J., Gnädig, N., Uhlemann, A.-C., Martin, R. E., Lehane, A. M., & Fidock, D. A. (2016). Globally prevalent PfMDR1 mutations modulate Plasmodium falciparum susceptibility to artemisinin-based combination therapies. Nature Communications, 7(1), 11553. https://doi.org/10.1038/ncomms11553

WHO. (2019). World Malaria Report. https://www.who.int/publications-detail/world-malaria-report-2019

Zhu, S. J., Hendry, J. A., Almagro-Garcia, J., Pearson, R. D., Amato, R., Miles, A., Weiss, D. J., Lucas, T. C., Nguyen, M., Gething, P. W., Kwiatkowski, D., McVean, G., & for the Pf3k Project. (2019). The origins and relatedness structure of mixed infections vary with local prevalence of P. falciparum malaria. ELife, 8. https://doi.org/10.7554/eLife.40845

