## Supplemental information for "Design and implementation of multiplexed amplicon sequencing panels to serve genomic epidemiology of infectious disease: a malaria case study"

#### Table of Contents:

| **Supplementary Figures** | |
|---|---|
| S1 Figure. AMPLseq panel composition in preliminary and final design stages. | Page 3 |
| S2 Figure. Amplicon sequence processing pipelines. | Page 4 |
| S3 Figure. AMPLseq read-pair counts from mock and clinical samples. | Page 5 |
| S4 Figure. Additional 4CAST and AMPLseq panel validation with mock mixed samples. | Page 6 |
| S5 Figure. Geographic attribution of combined simulated and empirical AMPLseq data. | Page 7 |
| S6 Figure. Assessing complexity of infection (COI) using AMPLseq. | Page 8 |
| S7 Figure. Comparison of COI signals from 4CAST, AMPLseq and WGS data types. | Page 9 |
| **Supplementary Notes** | |
| S1 Supporting information. | Page 10 |
| S2 Supporting information. Details and links to analysis pipelines. | Page 10 |
| S3 Supporting information. Note on two discarded AMPLseq microhaplotypes. | Page 12 |

| Supplementary Protocols | |
|---|---|
| S1 Protocol. 4CAST protocol. | Page 13 |
| S2 Protocol. AMPLseq protocol. | Page 15 |
| S3 Protocol. AMPure XP Bead Cleanup script for the KingFisher Flex. | Page 21 |
| S4 Protocol. gDNA extraction script for the KingFisher Flex. | Page 23 |

#### Supplementary Figures

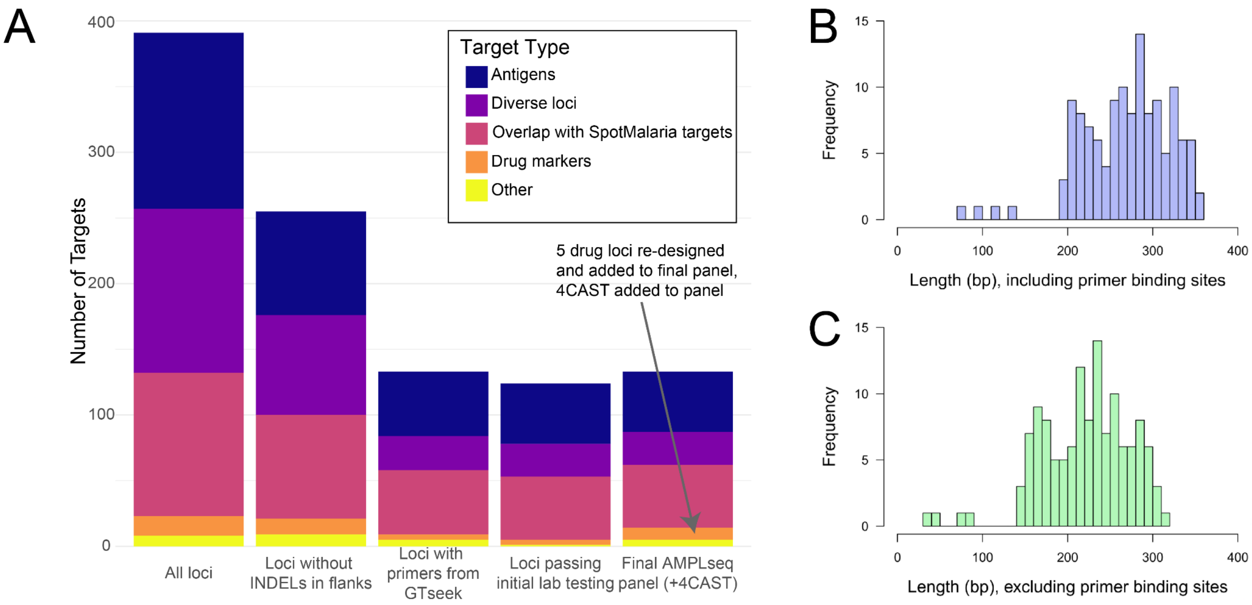

**S1 Figure. AMPLseq panel composition in preliminary and final design stages.**
A) Locus attribution during the design of the AMPLseq panel. Initial loci of interest were filtered out due to the presence of INDELs in flanking regions, primer interactions during the GTseek design process, and failure to amplify during lab testing, resulting in a final panel of 129 loci. Primers for several loci were redesigned after failure in initial lab testing and added back to the panel, resulting in an increase in loci between the last two columns. B) Amplicon length distribution of the final panel. Lengths range from 78 – 360 bp (median = 276 bp) including primer binding sites and C) from 35 – 320 bp (median = 226 bp) excluding primer binding sites.

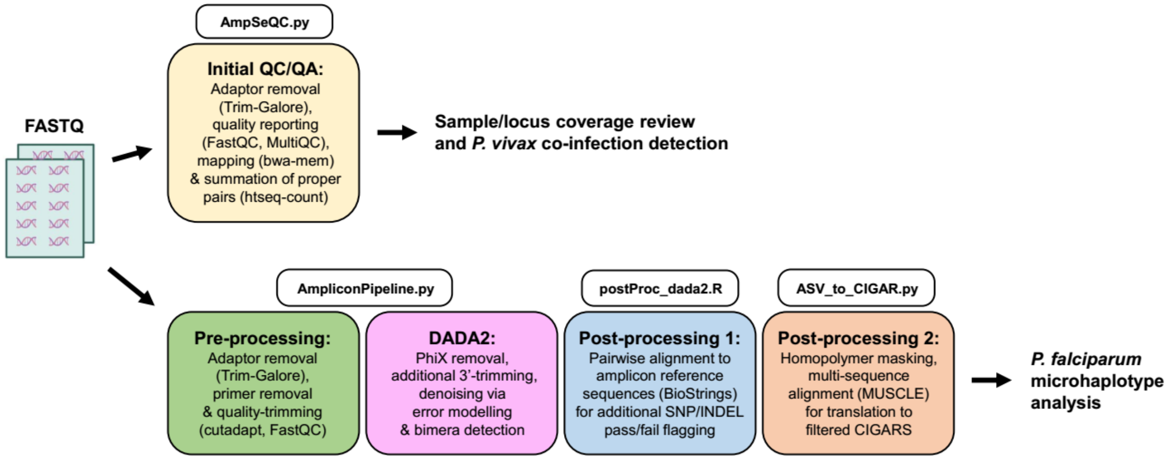

**S2 Figure. Amplicon sequence processing pipelines.**
Schematic of sequence processing pipelines used by 4CAST and AMPLseq. AmpSeQC provides preliminary read quality and quantity analyses (QC/QA) of both assays and enables *PvDHFR* detection by AMPLseq. The DADA2-based pipeline operates independently of AmpSeQC and represents the basis of P. falciparum microhaplotype analysis.

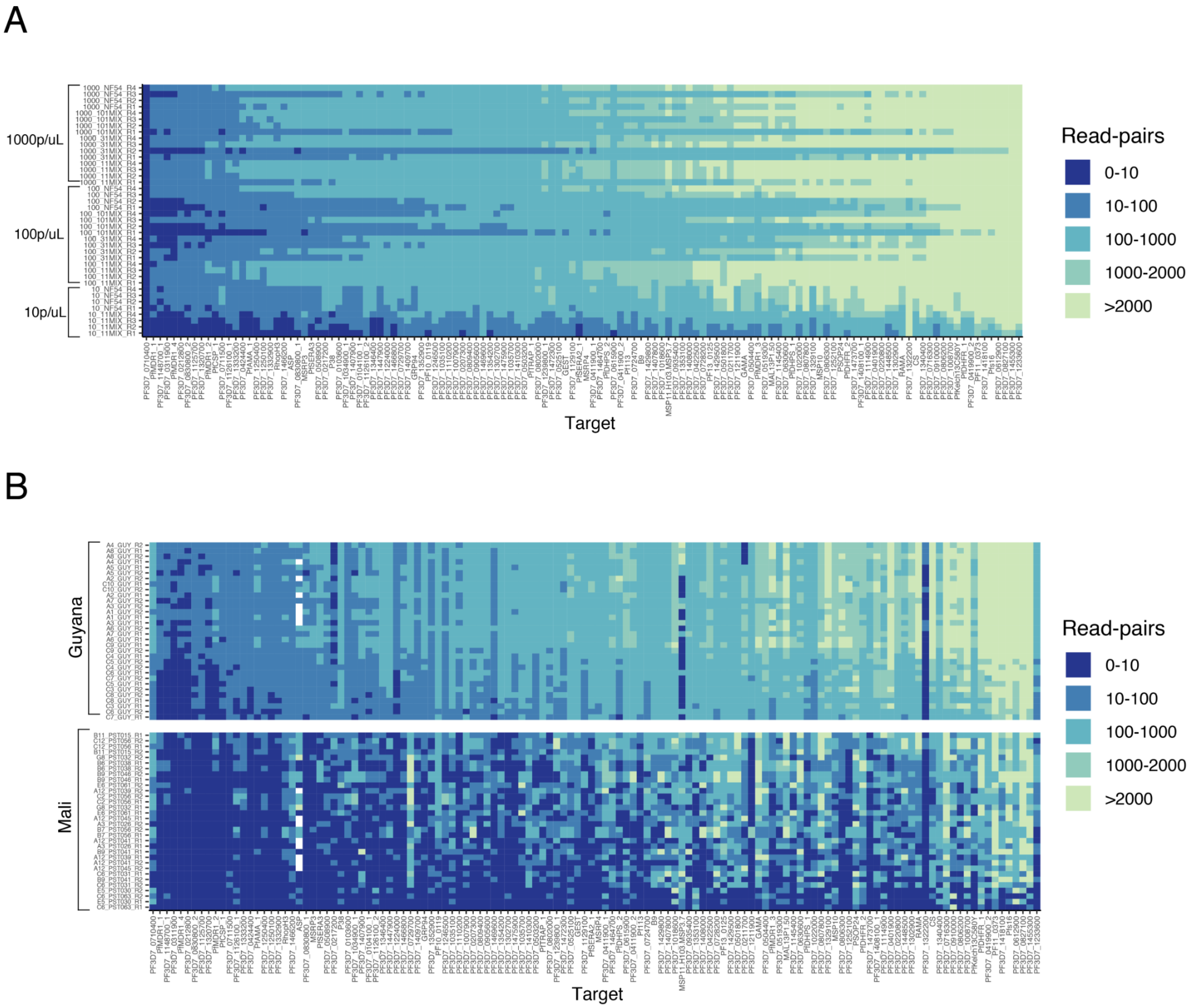

**S3 Figure. AMPLseq read-pair counts from mock and clinical samples.**
A) The heatmap shows the number of read-pairs obtained per mock sample (rows) at each AMPLseq locus (columns) from the DADA2-based pipeline output (see S2 Fig.). B) The same information is shown for the pre-amplified clinical sample sets.

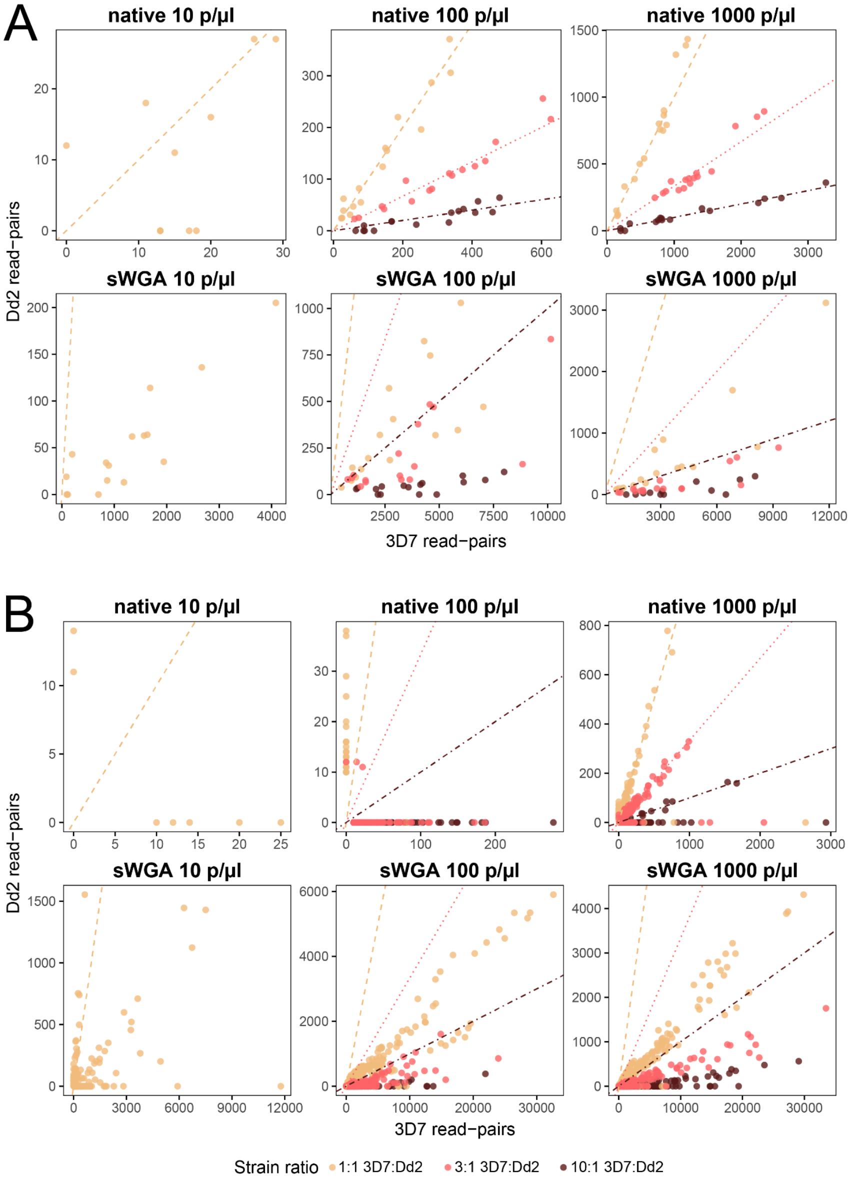

**S4 Figure. Additional 4CAST and AMPLseq panel validation with mock mixed samples.** A) Ratio of 4CAST read-pairs from microhaplotypes assigned to 3D7 (x-axis) or Dd2 (y-axis) from mock mixtures of these DNAs in ratios of 1:1 (tan), 3:1 (pink), and 10:1 (dark red) (Only the 1:1 ratio was tested at 10 p/µl). Dashed lines represent the expected ratio, and each point represents a 4CAST locus per sample (n=4 per condition). Data are shown from samples with 10, 100, or 1000 p/µl, and native DNA (top row) or sWGA (bottom row). (The native DNA at 1000 p/µl was shown in main Fig. 3C.) Read-pair ratios are very close to expected values in both native conditions, but sWGA distorts the read-pair ratios, greatly shifting the ratios towards 3D7. B) Ratio of AMPLseq read-pairs from the same templates as above, but with read-pairs from AMPLseq. (The native DNA at 1000 p/µl was shown in main Fig. 3D.) As with 4CAST, read-pair ratios are much more accurate on native DNA than sWGA DNA at 1000 p/µl. However, sWGA is recommended for AMPLseq samples with parasitemia of 100 p/µl or below, as the read-depth from native DNA at 100 p/µl is very low.

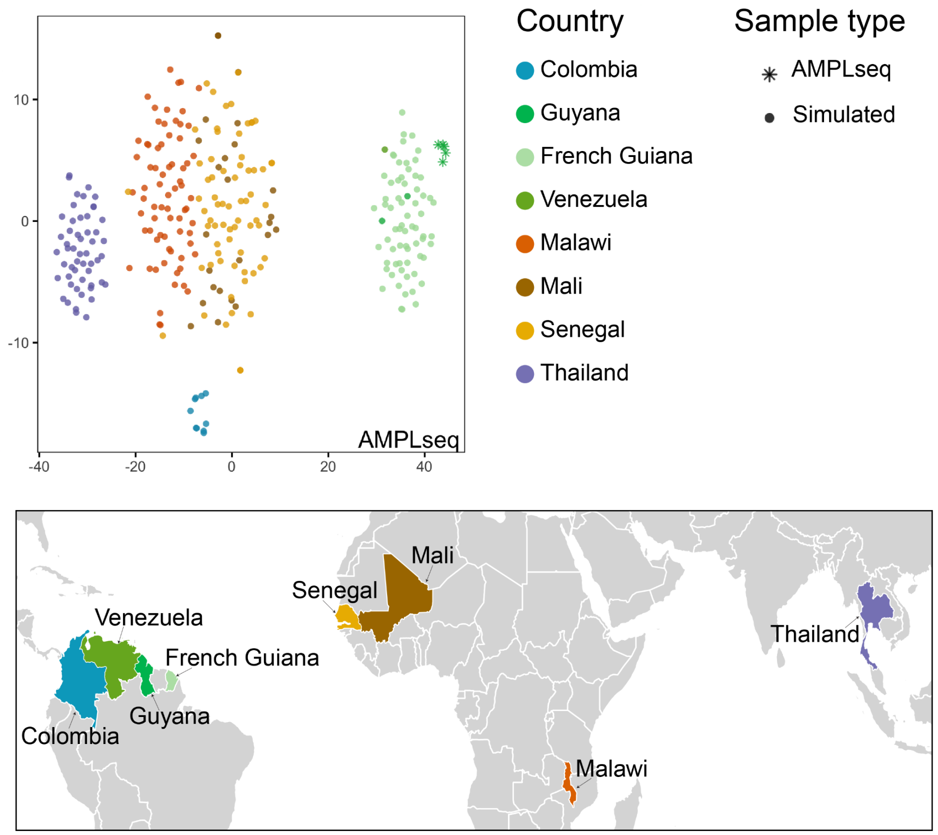

**S5 Figure. Geographic attribution of combined simulated and empirical AMPLseq data.** Visualization of WGS subset to AMPLseq loci (as in main Fig. 5A), but with the addition of empirical AMPLseq microhaplotypes, generated from samples from Guyana (n=5). After adding the empirical microhaplotypes, data were visualized using tSNE with the same parameters as in Fig. 5A. Samples are again colored by country, and the dots represent the data simulated from WGS, while the stars represent the empirical samples. The countries from which samples originate are colored in the map, for clarity of the geographic regions under consideration.

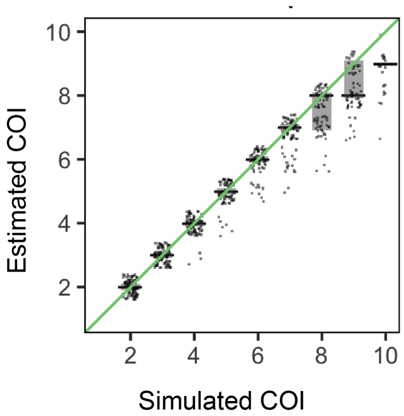

**S6 Figure. Assessing complexity of infection (COI) using AMPLseq.**
Scatter plots of estimated COI for samples simulated from combinations of monoclonal WGS data, subsetted to AMPLseq loci. The x-axis represents the number of monoclonal genomes combined into each simulation, and the y-axis represents the COI estimated using the simulated data. COI was simplistically estimated as the maximum number of unique microhaplotypes present at any locus per sample (n=100 samples per condition). Each dot represents a sample, jittered for visibility. The black bars represent the median, and the light grey boxes represent the 25th – 75th quantiles.

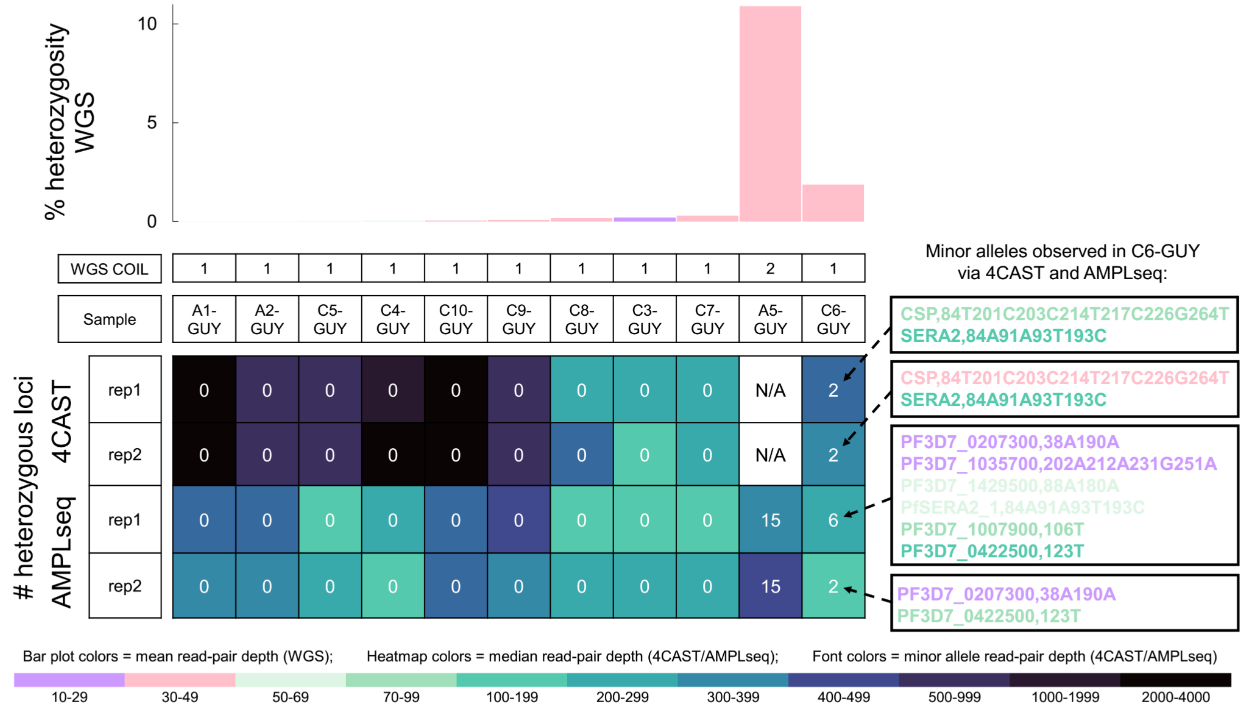

**S7 Figure. Comparison of COI signals from 4CAST, AMPLseq and WGS data types.** Multiallelic variant detection in ten Guyanese samples assayed by 4CAST (without sWGA), AMPLseq and WGS (both with sWGA). AMPLseq and WGS results for A5-GUY (the only Guyanese sample classified as COI>1 using The Real McCOIL (Chang et al., 2017) with WGS) are also included, but this sample was not assayed by 4CAST. While all three assays detect elevated multiallelic call rate in C6-GUY, the WGS signal (1.9% heterozygosity) is closer to baseline than to that in A5-GUY (10.9%), complicating conclusive COI classification. 4CAST and AMPLseq achieve significantly higher read-depth (see box fill colors) and detect two or more multiallelic loci in C6-GUY. WGS-based COI classification by The Real McCOIL is abbreviated as ‘WGS COIL’ below the bar plots of WGS heterozygosity rates.

### Supplementary Notes

**S1 Supporting information. Supplementary details of AMPLseq panel design.**

In designing the larger 'AMPLseq' multiplexed amplicon panel, we first built a large pool of candidate loci, anticipating significant attrition of candidates due to primer incompatibility. We deliberately targeted many coding sequences presumably subject to immune selection, as the primary intended analysis application (relatedness inference) is best supported by high diversity (Taylor et al., 2019). We used haplotypic diversity as a metric for identifying diverse loci. Haplotypic diversity is equivalent to heterozygosity for an outbred diploid (Nei & Tajima, 1981), and we use the term diversity here, unless referring to analyses by others where diversity is referred to as heterozygosity. We estimated diversity in 200 nucleotide (nt) sliding windows, spaced 50 nt apart, across the *P. falciparum* genome using the *scikit-allel* library (Miles et al., 2020). Specifically, we read in variant data (read_vcf function), masked positions with heterozygous variant calls (is_het and haploidify_samples functions), and estimated haplotype diversity (haplotype_diversity function). We made these estimates using Senegal WGS data from the Pf3k project (release 5; www.malariagen.net/projects/pf3k) and a collection of previously published French Guiana parasite data (Pelleau et al., 2015).

We next identified candidate loci for multiplexed amplicon design using these estimates of diversity. We selected the most diverse genomic window in antigens of interest, including previously published reactive antigens (Helb et al., 2015). To support relatedness inference, we also identified genomic windows within coding regions regardless of antigen status with haplotypic diversity > 0.7 in the Senegal data; we selected any window in a gene that we had not already included. Finally, we included SNPs in the SpotMalaria v1 SNP panel (Chang et al., 2019; Jacob et al., 2021) not already included in candidate genomic windows as a result of the other selection criteria.

We contracted the services of GTseek LLC (https://gtseek.com) to design multiplexed oligo panels using the Genotyping-in-Thousands by sequencing protocol (Campbell, Harmon, & Narum, 2015). In brief, we submitted a list of our candidate loci, with 100 nt flanks on either side of the selected genomic window. Using the Mali and Senegal data from the Pf3k project (MalariaGEN Plasmodium falciparum Community Project, 2016), we identified variants within these candidate loci. Candidate loci with insertions or deletions (INDELs) at ≥ 1% frequency in the flanking regions of the locus were culled to reduce potential for biased amplification, and any variants at ≥ 5% frequency in the locus were annotated. GTseek designed a set of primers with minimal cross-reactivity for a subset of our candidate loci.

We considered three loci for detection of *P. vivax* co-infection. *PvDHFR*, previously described in (Lefterova, Budvytiene, Sandlund, Färnert, & Banaei, 2015), was chosen because it outperformed multi-copy ribosomal subunit 18S (Rougemont et al., 2004) and subtelomeric repeat Pvr47 (Demas et al., 2011) amplification in multiplexed reaction.

**S2 Supporting information. Details and links to analysis pipelines.**

<u>AmpSeQC (https://github.com/broadinstitute/AmpSeQC)</u>

A general multiplexed amplicon sequencing and quality control pipeline specifically built for *Plasmodium falciparum* data. Input: paired-end fastq files, reference genome, gff3 annotation file of amplicon panel or genes. Output: tsv file of read counts per amplicon/gene per sample, MultiQC reports.

Within AmpSeQC, we processed demultiplexed paired-end FASTQ files with Trim-Galore (v0.6.6) (Krueger, 2016/2021) to remove Illumina sequencing adapter sequences and nucleotides with less than 20 PHRED base quality. We removed paired reads if trimming resulted in either read being shorter than 70 bp after trimming. We then aligned trimmed reads to the *P. falciparum* 3D7 genome (PlasmoDB v46) using BWA-MEM (v0.7.17-r1188) (Li, 2013) with the insert size parameters “-I 200,100,500,50”. We removed read-pairs that did not align with samtools (v1.11) (Danecek et al., 2021), and we removed read-pairs where either read had more than 5 bases clipped at the end with samclip (v0.4.0) (Seemann, 2018/2021). Additionally, we retained only properly paired reads with samtools (Danecek et al., 2021). We then reported read-pair counts per amplicon locus with htseq-count (v0.13.5) (Anders, Pyl, & Huber, 2015).

<u>Malaria amplicon pipeline (https://github.com/broadinstitute/malaria-amplicon-pipeline.git)</u>

An automated pipeline for processing of highly multiplexed amplicon sequencing reads that bundles together various pre-processing, QC and core denoising tools. It also includes a post processing component that filters processed de-noised sequences and converts them into pseudo-CIGAR variants. Primarily tested on amplicon data from *Plasmodium falciparum* genome. Input: paired-end fastq, reference target sequences, filter thresholds. Output: tsv file of pseudo-CIGAR variant read counts of given targets per sample, MultiQC reports.

We first used Trim-Galore (v0.6.6) (Krueger, 2016/2021) to remove Illumina sequencing adapters and cutadapt (v3.4) (Martin, 2011) to remove primer sequences from paired reads. We also removed read-pairs that did not contain expected pairs of forward and reverse primer sequences. Following these pre-processing steps outside of DADA2, we trimmed 2 bp from the 3’ ends of all reads to account for lower base quality generally observed in final cycles of sequencing. We also trimmed bases with PHRED scores below 5 from all affected 3’ ends. Further, we removed unidentified (N) bases and reads exceeding maximum expected error (maxEE) = 5. We ran the core denoising algorithm in SELF_CONSIST mode with MAX_CONSIST cycles set to 10 and OMEGA_A statistical evidence threshold set to $10^{-120}$. These non-default options were selected to promote consistent error modeling across diverse *P. falciparum* sample sets.

We mapped microhaplotypes obtained from DADA2 against a custom-built database of 3D7 and Dd2 reference sequences for each amplicon locus. We then recorded the edit distance (number of mismatching bases) of each mapped microhaplotype to 3D7, edit distance to Dd2, and length distance (net number of deleted or inserted bases) to 3D7. We flagged microhaplotypes with edit distances exceeding the maximum number of SNPs found among Pf3k samples (distinct cutoffs for each locus) and/or with length differences exceeding 10.5%, which represents the maximum length difference among corresponding 3D7 and Dd2 reference loci. Finally, we also flagged microhaplotypes identified as chimeras of sequences from two different loci. We used a custom R script to perform these DADA2 post-processing steps.

We summarized observed sequence polymorphism into a concise format by converting individual microhaplotypes into pseudo-CIGAR strings using a custom python script. In brief, we first discarded microhaplotypes flagged as bimeras or flagged for edit and/or length distances, as described above. We then generated a multi-sequence alignment for each locus using MUSCLE (v3.8.1551) (Edgar, 2004). Each multi-sequence alignment contained the 3D7 reference sequence (primer sequences removed) and the microhaplotypes mapped to a given locus. We used these alignments to mask homopolymer runs of five or more bases by alignment columns corresponding to these bases. To denote any single-nucleotide difference observed in a microhaplotype after these filtration steps, we used a character string starting with a number indicating the variant base position in the 3D7 reference locus followed by the variant base identity (alternate allele) at that position. We also denoted insertions starting first with reference locus position and then with ‘I=’ and the number of inserted bases. We denoted deletions by reference locus position, then ‘D=’ and the number of missing bases. If one or more resulting microhaplotypes within a sample had the same pseudo-CIGAR string (*e.g.*, due to differences only in homopolymer runs), we combined them by summing the read-pair counts for that sample across those microhaplotypes.

paneljudge (https://github.com/artaylor85/paneljudge)

An R package to judge the performance of a panel of genetic markers using simulated data. Given inter-marker distances and allele frequency estimates provided by the user, performance is judged using data (pairs of haploid genotypes) that are simulated under a hidden Markov model (HMM) (Taylor, Jacob, Neafsey, & Buckee, 2019) of relatedness between monoclonal malaria samples. To simulate data on a pair of haploid genotypes using paneljudge, the user must provide a vector of inter-locus distances, a matrix of loci allele frequencies, a relatedness parameter value, and a switch rate parameter value. Under the HMM of paneljudge, loci are considered categorical random variables whose realizations (alleles) are unordered (Taylor et al., 2019). Otherwise stated, under the HMM of paneljudge, loci are treated as point polymorphisms, thus ignoring the physical length and SNP distance differences between loci that contain multiple variants.

**S3 Supporting information. Note on two discarded AMPLseq microhaplotypes.**

Two microhaplotypes (pseudo-CIGAR strings) were discarded from AMPLseq analysis. The pseudo-CIGAR string PF3D7_1302900,1G was discarded because it occurred exclusively at multiallelic loci within high-concentration 3D7 and 3D7 + Dd2 mock samples (sWGA samples and native samples representing 10000 parasites/μl). The pseudo-CIGAR string PF3D7_0612900,215A was discarded because it occurred exclusively at heterozygous sites within 1:1 3D7 + Dd2 mock mixtures representing 100 parasites/μl. Prior to their removal, these two microhaplotypes were the only sources of false positivity (FP, see Methods) within the study's mock sample sets.

### Supplementary Protocols

#### S1 Protocol. 4CAST protocol.

<u>Reagents:</u>

1. KAPA HiFi 2X Master Mix (KK2600)
2. Nuclease-free (NF) H20
3. 4CAST oligonucleotides at 100uM (IDT), eluted in 1X TE or NF water*
4. IDT® for Illumina Nextera DNA Unique Dual Indexes (20027217)
5. *P. falciparum* genomic DNA

* 4CAST gene-specific primer with Illumina-compatible adapters (see Table S3 for sequences)

<u>Part 1</u>

***Primer aliquoting and first PCR should be prepared in a pre-PCR biosafety cabinet to avoid aerosol amplicon contamination***

<u>Making 4CAST primer cocktail</u>

1. In an eppendorf tube, combine 10 µL* of each primer at 100 µM following IDT instructions. Keep at -20 C.
2. For making working stock of 4CAST perform a 1/2 dilution (e.g., 80 µL primer mix + 80 µL nf water).
   *For over 96 samples this volume will have to be modified

<u>Master Mix for PCR1 for 1 sample. Load on a 96-well/384 plate.</u>

1. 5uL KAPA 2X HiFi Master Mix
2. 1.5uL of primer mix (at ~90nM/per primer)
3. 4 µL Sample (lower limit of detection is much lower but to start we would recommend 1 ng or more of *P. falciparum* DNA) (**or 4 µL of water for negative control**)

Place plate on thermalcycler and run for 10uL volume with following settings: **1.** 95.0° - 03:00, **2.** (98° - 00:20, 57° - 00:15, 62° - 00:30) **x 25 cycles**, **3.** 72° - 01:00, **4.** 4° - ∞

<u>Part 2</u>

***Aliquoting of Nextera-XT Indexes should be performed following Illumina instructions and in a pre-PCR biosafety cabinet to avoid aerosol amplicon contamination.* ***Setting up of PCR 2 should be performed in a post-PCR hood*******

<u>Master Mix for PCR 2 for 1 sample. Load on a 96-well plate.</u>

1. 5uL KAPA 2X HiFi Master Mix
2. 3uL of PCR1 (**or 3uL negative control from PCR1**)
3. 2uL nuclease-free water
4. 2.2uL of Nextera UDI primers (pre-aliquoted following Illumina instructions)

Place plate on thermalcycler and run for 10uL volume with following settings: **1.** 95.0° - 01:00, **2.** (95° - 00:15, 55° - 00:15, 72° - 00:30) **x 10 cycles**, **3.** 72° - 01:00, **4.** 4° - ∞

Sample pooling and PCR purification

1. Take ~8uL of each sample and mix in an eppendorf tube. If there are many samples, use a multichannel and combine into a reagent reservoir, mix well and add into an eppendorf tube.
2. AmpureXP bead cleanup (Beckman-Coulter, A63880)*
    a. Take 100 µL of combined sample into a PCR tube and add 55 µL of AmpureXP beads, mix well
    b. Incubate at room temperature for 5 minutes
    c. Place on magnetic rack for 5 minutes
    d. Remove the supernatant from the beads while still on the magnet and transfer into a new PCR tube.
    e. Remove supernatant from the magnetic stand and add 20 µL of beads. Mix well and incubate at room temperature for 5 minutes
    f. Place on magnetic rack for 3 minutes
    g. Discard supernatant and wash twice with 200 µL of **fresh** 80% Ethanol while still on the rack. Incubate for 30 seconds between the two washes.
    h. Remove tube from magnetic rack and allow remaining ethanol to evaporate (takes approximately 6-8 minutes, but keep an eye on the pellet- elute as/just before cracks begin to form)
    i. Elute in 30 µL Tris-HCl pH 8.0, incubate for 2 minutes, then place on magnetic rack and allow the pellet to form. If there are residual beads, pipette slightly under the total volume e.g., 25 µL
    **j. Add 2.5 Tris-HCl containing 1% Tween-20**

    **Note: Some of these times are variable based on the strength of the magnet.**

Confirmation of fragment size of 4CAST library and DNA quantification

1. DNA quantification can be performed using both an Agilent Bioanalyzer or a Qubit (high sensitivity kit)
2. Run bioanalyzer with DNA High Sensitivity protocol. Average fragment size should be between 400-500bp.

Sequencing

1. Dilute library to 4nM (MiSeq), or 200pM (iSeq)*
2. Add >10% of PhiX

** For dilution, use the concentration you get from qubit/bioanalyzer with the following formula:*
*(concentration in ng/uL)/(660g/mol)*(average library size in bp)*$10^6$ = concentration in nM*

#### S2 Protocol. AMPLseq protocol.

1) **Create primer cocktail stock:**

   a) Refer to Table S3 for the AMPLseq primer sequence information. Note that the majority of primers are stored in IDT plates at 200 uM, with well positions indicated in the spreadsheet. Exceptions are 4CAST primers (p96, p97, p253, p254, p255, p256, p257, p258) and selected DHPS/MDR1 primers (p576, p577, p578, p579, p580, p581, p582, p583, p584, p585); these are stored in tubes at 100 uM.

   b) Aliquot 2.68 ul of the following plate-primer wells [200 uM] into a single 2.0 ml Eppendorf tube. These 18 primers consistently overperform based on seq. results and are therefore aliquoted at this smaller volume (33% less than the input mode – see next step).

      p318 (M01)
      p328 (G02)
      p334 (M02)
      p376 (G05)
      p391 (F06)
      p397 (L06)
      p411 (J07)
      p416 (O07)
      p432 (O08)
      p451 (M01)
      p461 (G02)
      p467 (M02)
      p509 (G05)
      p524 (F06)
      p530 (L06)
      p544 (J07)
      p549 (O07)
      p565 (O08)

   c) Aliquot 4 uL each of the remaining 222 plate-primers [200 uM] into the same Eppendorf tube.

   d) Aliquot 8 uL each of the DHPS/MDR1 tube-primers [100 uM] (p576, p577, p578, p579, p580, p581, p582, p583, p584, p585) into the same Eppendorf tube.

   e) Aliquot 10.67 uL of each of the 4CAST tube-primers [100 uM] (p96, p97, p253, p254, p255, p256, p257, p258) into the same Eppendorf tube. These 8 primers consistently underperform based on seq. results and are therefore aliquoted at this larger volume (33% more than the input mode).

2) **Dilute primer cocktail stock to working concentration:**

   a) The primer pool stock tube has a total volume of 18 * 2.68 + 222 * 4 + 8 * 10.67 + 10 * 8 = 1101.6. The concentration mode per primer is therefore 4 ul * 200 uM / 1101.6 = 726 nM. We want to dilute it such that the concentration mode per primer is 200 nM.

      We should therefore dilute to ca. 27.5 %. We can do so by combining 400 ul primer pool with 1052 ul nuclease-free d$H_2O$ in a 2.0 ml Eppendorf tube.

b) Divide the diluted tube into five 300 ul aliquots and label clearly (work. conc.) so as not to confuse with remaining primer pool stock. Store stock and work. conc. aliquots at -20 °C.

3) **PCR1:**

a) Prepare PCR1 cocktail. The following volumes are needed per sample. Multiply these by the number of samples (plus 3 extra samples to accommodate pipetting error):

5 ul Qiagen Plus Master Mix (2x)
1.5 ul primer cocktail [200 nM per primer working conc.]
0.5 ul of nuclease-free $dH_2O$

b) Add 7 ul PCR1 cocktail to every plate well.

c) Add 3 ul of sample genomic DNA (or negative/positive control templates). It is best to place controls randomly throughout plate.

d) Place plate on a thermocycler with the following amplification settings:

1x
95 °C – 15:00

5x
95 °C – 00:30
57 °C – 00:30 (5% ramp; ~0.3° per second)
72 °C – 02:00

20x
95 °C – 00:30
65 °C – 00:30
72 °C – 00:30

1x
4° C - hold

4) **PCR1 product dilution:**

a) Create 1/13 PCR1 product dilution by transferring 2 ul of each product into a new plate and diluting with 24 ul nuclease-free $dH_2O$.

5) **PCR2:**

a) Prepare PCR2 cocktail. The following volumes are needed per sample. Multiply these by the number of samples (plus 3 extra samples to accommodate pipetting error):

5 ul KAPA HiFi HotStart ReadyMix (2x)
2.2 ul unique dual index (from 10 uM plate; we use IDT for Illumina – Nextera DNA UD Index Sets A-D; it is best to use distinct index sets when performing sequential seq. runs – e.g., alternate between sets A + B and C + D)

b) Add 7.2 ul PCR2 cocktail to every plate well.

c) Add 3 ul diluted PCR1 product.

d) Place plate on a thermocycler with the following amplification settings:

1x
95 °C – 03:00

10x
98 °C – 00:20
65 °C – 00:30
72 °C – 00:30

1x
72° C – 01:00

1x
4° C – hold

6) **SequalPrep normalization:**

***This normalization step is OPTIONAL (you can proceed directly to step 7). It was too aggressive in our hands, so it is only recommended if using DNA extracts representing >>1000 parasites/ul and when sample read depth balance is critical to the study question.

a) Transfer 10 ul PCR2 product to SequalPrep Normalization Plate. Some wells may have less than 10 ul due to evaporation – proceed anyway.

b) Add 10 ul SequalPrep Normalization Binding Buffer, pipette mix thoroughly and let incubate for one hour at room temperature.

c) Without scraping the side of wells, aspirate liquid and discard.

d) Dispense 50 ul SequalPrep Normalization Wash Buffer to every well. Mix by pipetting up and down twice and completely aspirate the buffer from the wells and discard. You many need to invert and tap the plate on paper towels in order to remove the residual wash buffer from the wells. A small amount of wash buffer (1 – 3 ul) is typical and does not affect downstream applications.

e) Add 20 ul SequalPrep Normalization Elution Buffer to each well. Seal, vortex, and briefly centrifuge the plate. Incubate at room temperature for 5 minutes.

f) Take 10 ul from each well of the normalized plate and combine into a 1.5 ml Eppendorf tube (you can use a boat or a PCR tube strip to enable multi-channel pipetting). Store normalized PCR2 product remainders (ca. 10 ul each) at -20 °C.

7) **Combine PCR2 products:**

***If SequalPrep normalization was performed, then ignore this step and proceed directly to step 8 because you have already combined normalized products.

a) Combine 4 ul of each PCR2 product into a 1.5 ml Eppendorf tube. Store PCR2 product remainders (ca. 6 ul each) at -20 °C.

8) **AMpure XP bead size selection (left-tailed clean-up):**

***Equilibrate beads to room temperature for 30 minutes prior to using them. It is also useful to take BioAnalyzer reagents out of 4 °C at this time such that you can proceed to QC directly after size selection.

a) Aliquot 60 ul of combined PCR2 product into each of five tubes within a PCR tube strip. Each of these five aliquots will receive a different AMpure bead input volume (0.9x = 54 ul, 0.8x = 48 ul, 0.7x = 42 ul, 0.6x = 36 ul, 0.5x = 30 ul) so that we can select the clean-up with the best QC result for sequencing. The 0.8x clean-up usually works quite well, but variation is common based on PCR2 product concentrations and pipetting error.

b) Vortex AMpure beads thoroughly immediately prior to use. Add 54 ul, 48 ul, 42 ul, 36 ul, and 30 ul to the five sample aliquots, respectively. Mix thoroughly.

c) Incubate at room temperature for 5 minutes.

d) Place on magnetic stand until solution clears (3+ minutes).

e) Discard the supernatant without disturbing the bead pellet.

f) While still on the magnetic stand, add 200 ul of fresh 80% ethanol to the beads and incubate for 30 seconds. Remove supernatant and discard. Repeat this wash once.

g) While still on the magnetic stand, remove any residual ethanol with a small (e.g., p20) pipette and/or allow ethanol to evaporate for 2 minutes (with tubes uncovered). Do not exceed 2 minutes evaporation time.

h) Remove from magnetic stand and add 15.5 ul EB buffer (10 mM Tris-Cl, pH 8.5). Mix thoroughly and let incubate at room temperature for 5 minutes.

i) Place on magnetic stand until solution clears (3+ minutes).

j) Collect 15 ul supernatant without disturbing the pellet.

**9) BioAnalyzer QC / library selection for sequencing:**

***Ensure reagents have equilibrated to room temperature for 30 minutes.

a) It is best to first verify via Qubit fluorometer that your input concentrations fall within BioAnalyzer's linear dynamic range (ca. 50 pg/ul – 10 ng/ul). In our experience, size-selected libraries generally require 1:100 dilution in EB buffer (10 mM Tris-Cl, pH 8.5) + 0.1% Tween-20 to reach appropriate concentrations for BioAnalyzer.

b) Follow manufacturer's instructions to run the BioAnalyzer. An example of results is shown below. In this case the 0.8x bead clean-up is chosen for sequencing because it contains no small fragments (primer polymers that can compromise efficient sequencing) and the target peak range is still fully intact.

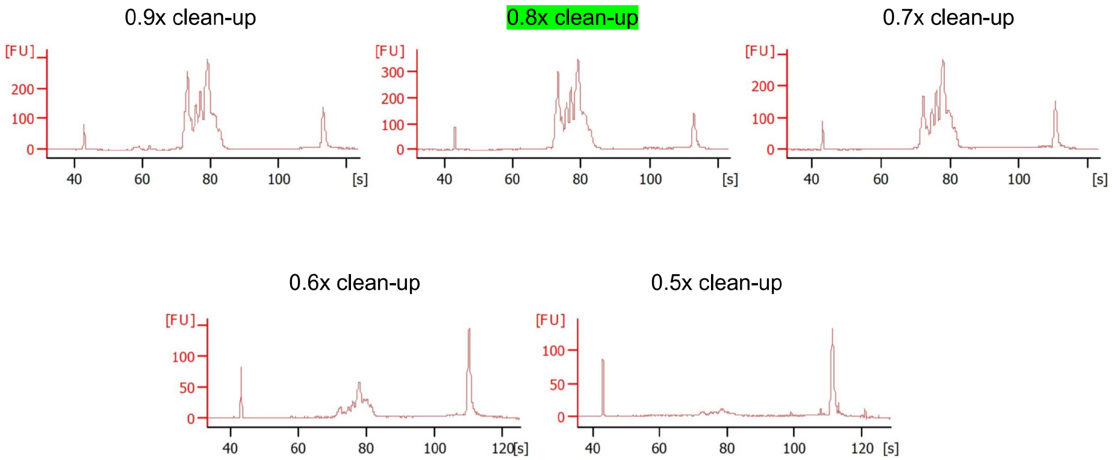

#### 10) Final library quantification and sequencing input preparation:

***Accurate quantification is essential to avoid under- or overclustering of the flow cell. It is useful to triangulate results from Qubit, BioAnalyzer and qPCR methods. You can convert the ng/ul values reported by Qubit to nM values with the following formula:

($10^6$ * conc. in ng/uL) / (660 g/mol * average library size in bp) = conc. in nM

It is however not uncommon for quantifications from Qubit, BioAnalyzer and qPCR to differ substantially. The results of qPCR should be considered the most reliable because qPCR can specifically quantify inserts complete with adaptors as opposed to total DNA.

a) We use KAPA Library Quantification Kit for Illumina platforms following the manufacturer's instructions. We include the optional S0 standard (diluted 1:10) for additional method control. The 1:100 library dilution used previously for BioAnalyzer generally needs another 1:1000 dilution to fall within the qPCR kit's standard curve (0.002 pM – 20 pM). We generally also use the qPCR to verify the concentration of PhiX to be used in sequencing. The PhiX qPCR input is a 1:1000 dilution of the 10 nM stock tube purchased from Illumina. Furthermore, it is useful to include previously sequenced libraries (diluted appropriately) in the qPCR for reference.

b) Following the qPCR, you can calibrate the quantification results for your library based on average fragment length measured previously via BioAnalyzer.

c) Dilute library to 4 nM (MiSeq) or 200 pM (iSeq). Follow the corresponding sequencing guides:

MiSeq:
https://support.illumina.com/content/dam/illumina-support/documents/documentation/system_documentation/miseq/miseq-denature-dilute-libraries-guide-15039740-10.pdf (Protocol A)

iSeq:
https://support.illumina.com/content/dam/illumina-support/documents/documentation/system_documentation/iseq100/iseq-100-system-guide-1000000036024-07.pdf

Optimally balancing the trade-off between output quantity and quality is absolutely key to maximizing target coverage. Library and PhiX flowcell loading concentrations play a critical part in achieving this balance.

While very high quality can be achieved by underclustering the flowcell and/or oversupplying PhiX, target yield will be low. Conversely, lots of target data can be produced by overclustering the flowcell and/or undersupplying PhiX, but error rates will be high.

We generally find 6 – 8 % PhiX sufficient for raising GC content in GTseq. We recommend not to exceed 10% PhiX (in contrast to 4CAST, which seems to require closer to 25% PhiX).

We aim for 800 – 1000 K/mm$^2$ cluster density using MiSeq Reagent Kit v2 (2 x 250 bp) despite Illumina's recommendations for 1000 – 1200 K/mm$^2$. The latter may only be appropriate for more base-balanced libraries.

In our experience, a good AMPLseq run is one with > 90% clusters passing filter (PF), > 90% average Q30 score and > 8 Gbp yield.

**S3 Protocol. AMPure XP bead clean-up script for the KingFisher Flex.**
The script is summarized via ThermoFisher Scientific BindIt 4.1 software export of protocol steps. ‘Sample-Plate’ wells contain 50 µl sample and 90 µl AMPure XP beads. ‘Wash-Plate-1’ and ‘Wash-Plate-2’ wells contain 200 µl freshly prepared 80% ethanol. ‘Elution-Plate’ wells contain 50 µl low TE buffer (10 mM Tris-HCl (pH 8.0) + 0.1 mM EDTA).

| | | |
|---|---|---|
| **Tip1** | **96 DW Tip-Comb** | |
| **Pick-Up** | **Tip-Comb** | |
| **Binding** | **Sample-Plate** | |
| Beginning of step | Precollect | No |
| | Release beads | Yes |
| Mixing / heating: | Mixing time, speed | 00:10:00, Medium |
| | Heating during mixing | No |
| End of step | Postmix | No |
| | Collect count | 5 |
| | Collect time [s] | 1 |
| **CollectBeads1** | **Sample-Plate** | |
| | Collect count | 5 |
| | Collect time [s] | 1 |
| **CollectBeads2** | **Sample-Plate** | |
| | Collect count | 5 |
| | Collect time [s] | 1 |
| **Wash1** | **Wash-Plate-1** | |
| Beginning of step | Precollect | No |
| | Release beads | No |
| Mixing / heating: | Mixing time, speed | 00:00:15, Medium |
| | Heating during mixing | No |
| End of step | Postmix | No |
| | Collect beads | No |
| **Wash2** | **Wash-Plate-2** | |
| Beginning of step | Precollect | No |
| | Release beads | No |
| Mixing / heating: | Mixing time, speed | 00:00:15, Medium |
| | Heating during mixing | No |
| End of step | Postmix | No |
| | Collect beads | No |
| **Dry1** | **Wash-Plate-2** | |

| | | |
|---|---|---|
| | Dry time | 00:02:00 |
| | Tip position | Outside well / tube |
| Elute | Elution-Plate | |
| Beginning of step | Precollect | No |
| | Release beads | Yes |
| Mixing / heating: | Mixing time, speed | 00:05:00, Medium |
| | Heating during mixing | No |
| End of step | Postmix | No |
| | Collect beads | No |
| Remove beads | Elution-Plate | |
| Beginning of step | Precollect | Yes |
| | Release beads | No |
| Mixing / heating: | Mixing time, speed | 00:01:00, Slow |
| | Heating during mixing | No |
| End of step | Postmix | No |
| | Collect count | 5 |
| | Collect time [s] | 30 |
| Leave | Wash-Plate-1 | |

**S4 Protocol. Genomic DNA extraction script for the KingFisher Flex.**
The script is summarized via ThermoFisher Scientific BindIt 4.1 software export of protocol steps. ‘Sample-Plate’ wells contain 480 µl Proteinase K Mix (400 µl nuclease-free $H_2O$, 40 µl Proteinase K and 40 µl Enhancer Solution previously incubated with the dried blood spot sample) and 400 µl Binding Solution provided by KingFisher Flex-Ready DNA Ultra 2.0 Prefilled Plates. All other plates used by the protocol come prefilled within this kit and are centrifuged before use.

| | | |
|---|---|---|
| Tip1 | 96 DW Tip-Comb | |
| Pick-Up | Tip-Comb | |
| Pick up beads | Bead-Plate | |
| | Collect count | 3 |
| | Collect time [s] | 3 |
| Binding | Sample-Plate | |
| Beginning of step | Precollect | No |
| | Release beads | Yes |
| Mixing / heating: | Mixing time, speed | 00:05:00, Fast |
| | Heating during mixing | No |
| End of step | Postmix | No |
| | Collect count | 5 |
| | Collect time [s] | 0 |
| CollectBeads1 | Sample-Plate | |
| | Collect count | 5 |
| | Collect time [s] | 0 |
| Wash I | Wash-I-Plate | |
| Beginning of step | Precollect | No |
| | Release time, speed | 00:00:20, Bottom mix |
| Mixing / heating: | Shake 1 time, speed | 00:00:10, Bottom mix |
| | Shake 2 time, speed | 00:00:10, Fast |
| | Loop count | 3 |
| | Heating during mixing | No |
| End of step | Postmix | No |
| | Collect count | 5 |
| | Collect time [s] | 0 |
| Wash II_1 | Wash-II-1-Plate | |
| Beginning of step | Precollect | No |
| | Release time, speed | 00:00:20, Fast |

| | | |
|---|---|---|
| Mixing / heating: | Shake 1 time, speed | 00:00:10, Bottom mix |
| | Shake 2 time, speed | 00:00:10, Fast |
| | Loop count | 2 |
| | Heating during mixing | No |
| End of step | Postmix | No |
| | Collect count | 4 |
| | Collect time [s] | 1 |

| Wash II_2 | Wash-II-2-Plate | |
|---|---|---|
| Beginning of step | Precollect | No |
| | Release beads | Yes |
| Mixing / heating: | Mixing time, speed | 00:00:30, Fast |
| | Heating during mixing | No |
| End of step | Postmix | No |
| | Collect count | 4 |
| | Collect time [s] | 1 |

| Dry | Wash-II-2-Plate | |
|---|---|---|
| | Dry time | 00:02:00 |
| | Tip position | Outside well / tube |

| Elution | Elution-Plate | |
|---|---|---|
| Beginning of step | Precollect | No |
| | Release beads | Yes |
| Mixing / heating: | Shake 1 time, speed | 00:00:15, Bottom mix |
| | Shake 2 time, speed | 00:00:45, Medium |
| | Loop count | 6 |
| | Heating temperature [°C] | 75 |
| | Preheat | Yes |
| End of step | Postmix | No |
| | Collect count | 1 |
| | Collect time [s] | 0 |

| Collect beads | Elution-Plate | |
|---|---|---|
| Beginning of step | Precollect | No |
| | Release beads | No |
| Mixing / heating: | Mixing time, speed | 00:02:00, Slow |
| | Heating during mixing | No |
| End of step | Postmix | No |
| | Collect beads | No |

| Leave | Tip-Comb |
|---|---|

#### References

Anders, S., Pyl, P. T., & Huber, W. (2015). HTSeq—a Python framework to work with high-throughput sequencing data. *Bioinformatics*, *31*(2), 166–169. https://doi.org/10.1093/bioinformatics/btu638

Campbell, N. R., Harmon, S. A., & Narum, S. R. (2015). Genotyping-in-Thousands by sequencing (GT-seq): A cost effective SNP genotyping method based on custom amplicon sequencing. *Molecular Ecology Resources*, *15*(4), 855–867. https://doi.org/10.1111/1755-0998.12357

Chang, H.-H., Worby, C. J., Yeka, A., Nankabirwa, J., Kamya, M. R., Staedke, S. G., Dorsey, G., Murphy, M., Neafsey, D. E., Jeffreys, A. E., Hubbart, C., Rockett, K. A., Amato, R., Kwiatkowski, D. P., Buckee, C. O., & Greenhouse, B. (2017). THE REAL McCOIL: A method for the concurrent estimation of the complexity of infection and SNP allele frequency for malaria parasites. *PLoS Computational Biology*, *13*(1), e1005348. https://doi.org/10.1371/journal.pcbi.1005348

Danecek, P., Bonfield, J. K., Liddle, J., Marshall, J., Ohan, V., Pollard, M. O., Whitwham, A., Keane, T., McCarthy, S. A., Davies, R. M., & Li, H. (2021). Twelve years of SAMtools and BCFtools. *GigaScience*, *10*(2). https://doi.org/10.1093/gigascience/giab008

Demas, A., Oberstaller, J., DeBarry, J., Lucchi, N. W., Srinivasamoorthy, G., Sumari, D., Kabanywanyi, A. M., Villegas, L., Escalante, A. A., Kachur, S. P., Barnwell, J. W., Peterson, D. S., Udhayakumar, V., & Kissinger, J. C. (2011). Applied genomics: data mining reveals species-specific malaria diagnostic targets more sensitive than 18S rRNA. *Journal of Clinical Microbiology*, *49*(7), 2411–2418. https://doi.org/10.1128/JCM.02603-10

Edgar, R. C. (2004). MUSCLE: multiple sequence alignment with high accuracy and high throughput. *Nucleic Acids Research*, *32*(5), 1792–1797. https://doi.org/10.1093/nar/gkh340

Krueger, F. (2021). *FelixKrueger/TrimGalore* [Perl]. https://github.com/FelixKrueger/TrimGalore (Original work published 2016)

Lefterova, M. I., Budvytiene, I., Sandlund, J., Färnert, A., & Banaei, N. (2015). Simple real-time PCR and amplicon sequencing method for identification of plasmodium species in human whole blood. *Journal of Clinical Microbiology*, *53*(7), 2251–2257. https://doi.org/10.1128/JCM.00542-15

Li, H. (2013). Aligning sequence reads, clone sequences and assembly contigs with BWA-MEM. *ArXiv:1303.3997 [Preprint]*. http://arxiv.org/abs/1303.3997

MalariaGEN Plasmodium falciparum Community Project. (2016). *The Pf3K Project: pilot data release 5.* www.malariagen.net/data/pf3k-5.

Martin, M. (2011). Cutadapt removes adapter sequences from high-throughput sequencing reads. *EMBnet.Journal*, *17*(1), 10–12. https://doi.org/10.14806/ej.17.1.200

Rougemont, M., Van Saanen, M., Sahli, R., Hinrikson, H. P., Bille, J., & Jaton, K. (2004). Detection of Four Plasmodium Species in Blood from Humans by 18S rRNA Gene Subunit-Based and Species-Specific Real-Time PCR Assays. *Journal of Clinical Microbiology*, *42*(12), 5636–5643. https://doi.org/10.1128/JCM.42.12.5636-5643.2004

Seemann, T. (2021). *samclip* [Perl]. https://github.com/tseemann/samclip (Original work published 2018)

Taylor, A. R., Jacob, P. E., Neafsey, D. E., & Buckee, C. O. (2019). Estimating Relatedness Between Malaria Parasites. *Genetics*, *212*, 1337–1351.
